# Exploiting the Reverse Vaccinology Approach to Design Novel Subunit Vaccine against Ebola Virus

**DOI:** 10.1101/2020.01.02.20016311

**Authors:** Md. Asad Ullah, Bishajit Sarkar, Syed Sajidul Islam

**Affiliations:** Department of Biotechnology and Genetic Engineering, Faculty of Biological Sciences, Jahangirnagar University, Savar, Dhaka, Bangladesh

**Keywords:** Ebola virus, vaccine, immunoinformatics, bioinformatics, epitopes

## Abstract

Ebola virus is a highly pathogenic RNA virus that causes haemorrhagic fever in human. With very high mortality rate, Ebola virus is considered as one of the dangerous viruses in the world. Although, the Ebola outbreaks claimed many lives in the past, no satisfactory treatment or vaccine have been discovered yet to fight against Ebola. For this reason, in this study, various tools of bioinformatics and immunoinformatics were used to design possible vaccines against Zaire Ebola virus strain Mayinga-76. To construct the vaccine, three potential antigenic proteins of the virus, matrix protein VP40, envelope glycoprotein and nucleoprotein were selected against which the vaccines would be designed. The MHC class-I, MHC class-II and B-cell epitopes were determined and after robust analysis through various tools and molecular docking analysis, three vaccine candidates, designated as EV-1, EV-2 and EV-3, were constructed. Since the highly conserved epitopes were used for vaccine construction, these vaccine constructs are also expected to be effective on other strains of Ebola virus like strain Gabon-94 and Kikwit-95. Next, the molecular docking study on these vaccine constructs were analyzed by molecular docking study and EV-1 emerged as the best vaccine construct. Later, molecular dynamics simulation study revealed the good performances as well as good stability of the vaccine protein. Finally, codon adaptation and in silico cloning were conducted to design a possible plasmid (pET-19b plasmid vector was used) for large scale, industrial production of the EV-1 vaccine.

## 1. Introduction

Highly pathogenic Ebola viruses are non-segmented RNA viruses with high mortality rates that are a distinguishing feature of this human pathogen associated with Ebola haemorrhagic fever [1, 2]. The first recorded Ebola outbreaks were observed in Africa continent between June and November of 1976 and 53% (150 of 284 victims) was the mortality rate [3, 4]. Currently, there is no effective anti-viral treatment for the rapid progression of Ebola that allows little opportunity to develop natural immunity [5]. Ebola virus was subsequently defined as the prototype viruses of a new taxonomic family, filoviridae [6]. Ebola viruses are subdivided into four different subtypes, Zaire, Sudan, Cote d’Ivoire, and Reston. The latest discovery includes fourth African species of human-pathogenic ebola virus is the Bundibugyo Ebola virus strain. Beside of this, there are many more pathogenic strains includes Zaire Mayinga-76, Gabon-96, Kikwit-95, Eckron-76 etc. [7, 8, 9]. Among them Zaire Mayinga-76 strain and its vaccination study is performed in this in silico research article. Ebola hemorrhagic viral fever is a severe, often deadly disease of humans and nonhuman primates cause by non-segmented, single stranded RNA molecules of Ebola virus [10]. After the attack of Ebola virus, the immune system causes a systemic inflammatory response that causes the impairment of the vascular, coagulation, asthenia and arthralgia systems, which leads to multi-organ failure, resembling septic shock are some of the indicators of ebola virus infections [7]. In 2014, World Health Organization (WHO) organized an emergency meeting attended by 60 researchers and public health administrators to assess and produce safe and effective Ebola vaccines as soon as possible [11]. A recent study shows that a trial vaccine called recombinant vesicular stomatitis virus–Zaire Ebola virus (rVSV-ZEBOV), which has shown to be safe and protective against the Zaire strain is recommended for use in Ebola outbreaks [12]. However, despite a huge number of research and investigation, there is no effective and satisfactory therapeutic clarification or vaccination has been invented so far against the Mayinga-76 strain of Ebola virus. Even if an effective and safe vaccine can be produced, it is not likely to be hundred percent effective to succeed in slowing down and stopping the current outbreak [13]. Development of an effective and safe Ebola virus vaccine has been hindered by a lack of knowledge. In this study, an effort has been made to develop an effective Ebola vaccine using various tools of bioinformatics, immunoinformatics and reverse vaccinology (**Figure 01**). Reverse vaccinology is a process of vaccine development where the novel antigens are identified by analyzing the genomic information of an organism or a virus. In reverse vaccinology, various tools of in silico biology are used to discover the novel antigens by studying the genetic makeup of a pathogen and the genes that could lead to good epitopes are determined. This method is a quick easy and cost-effective way to design vaccine. Reverse vaccinology is successfully used for designing vaccines for many viruses like zika virus, chikungunya virus etc. [14].

**Figure 01.**
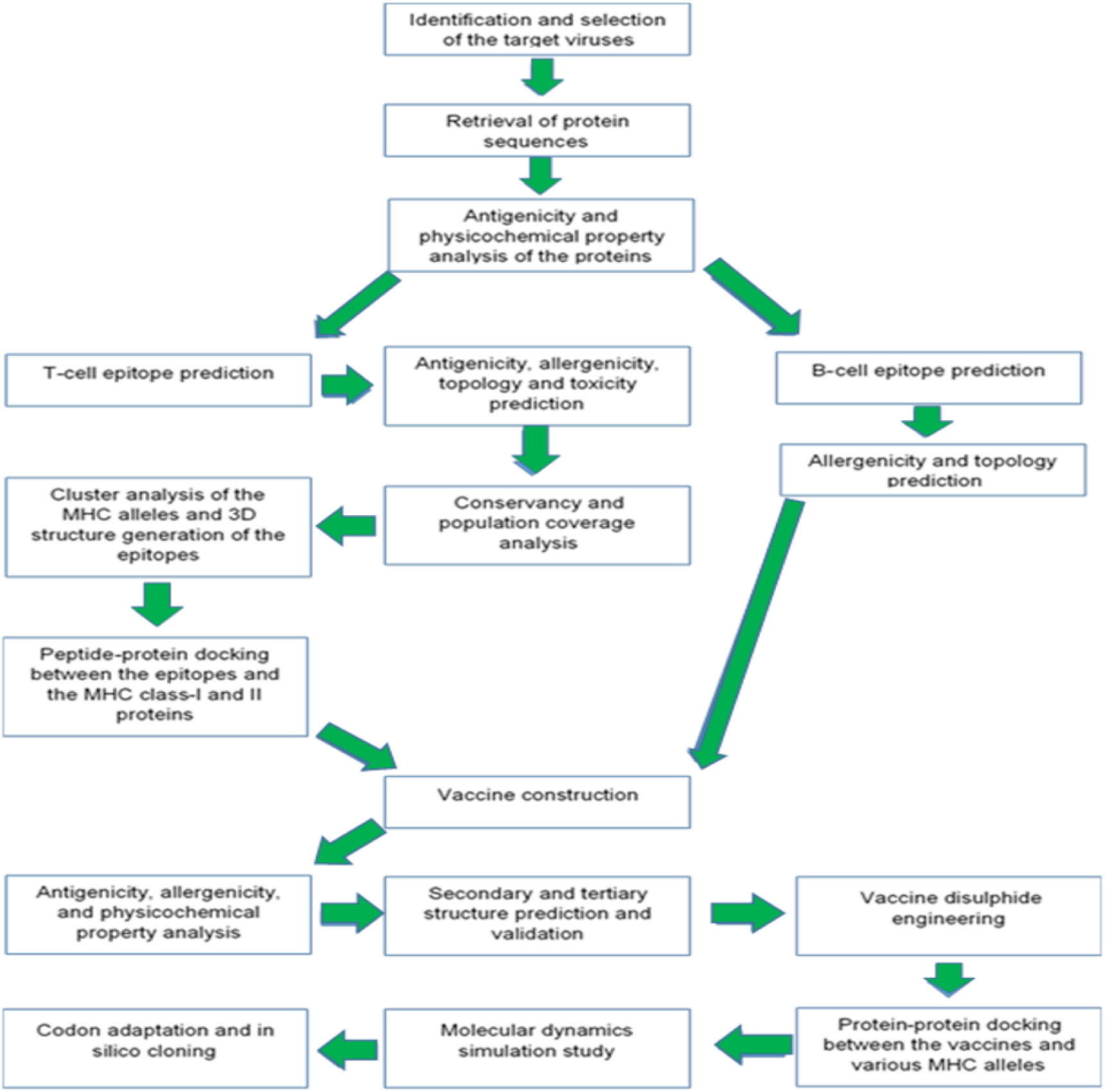
The flowchart of the procedures carried out in conducting the vaccine designing experiment.

## 2. Materials and methods

The current experiment focuses on development of vaccines against the [Zaire Ebola virus (strain Mayinga-76)](https://www.uniprot.org/taxonomy/128952).

### 2.1. Strain Identification and Selection

The strain of the Ebola virus strain Mayinga-76 was identified and selected by analyzing different entries of the online server of National Center for Biotechnology Information or NCBI (https://www.ncbi.nlm.nih.gov/).

### 2.2. Retrieval of the Protein Sequences

The viral envelope glycoprotein (accession number: Q05320), matrix protein VP40 (accession number: [Q05128](https://www.uniprot.org/uniprot/Q05320)) and nucleoprotein (accession number: P18272) were retrieved from the UniProt Knowledgebase (UniProtKB) tool of the online server UniProt (https://www.uniprot.org/).

### 2.3. Antigenicity Prediction and Physicochemical Property Analysis of the Protein Sequences

The antigenicity of the protein sequences were predicted by online server for antigenicity analysis, VaxiJen v2.0 (http://www.ddg-pharmfac.net/vaxijen/VaxiJen/VaxiJen.htm), keeping the threshold at 0.4 [15, 16, 17]. The various physicochemical properties of the selected protein sequences were determined by ExPASy’s online tool ProtParam (https://web.expasy.org/protparam/) [18].

### 2.4. T-cell and B-cell Epitope Prediction

The T-cell and B-cell epitopes of the selected protein sequences were predicted using online epitope prediction server Immune Epitope Database or IEDB (https://www.iedb.org/). The IEDB database contains huge amount of experimental data on T-cell epitopes and antibodies. These data are collected from various experiments that are carried on human, non-human primates and other animals. It is a server that allows robust analysis on various epitopes in the context of some tools: population coverage, conservation across antigens and clusters with similar sequences [19]. The MHC class-I restricted CD8+ cytotoxic T-lymphocyte (CTL) epitopes of the selected sequences were obtained using NetMHCpan EL 4.0 prediction method for HLA-A*11-01 allele. The MHC class-II restricted CD4+ helper T-lymphocyte (HTL) epitopes were obtained for HLA DRB1*04-01 allele using Sturniolo prediction method. Ten of the top twenty MHC class-I and MHC class-II epitopes were randomly selected based on their percentile scores and antigenicity scores (AS). Five random B-cell lymphocyte epitopes (BCL) were selected based on their length (the sequences that have ten amino acids or above were selected) and obtained using Bipipered linear epitope prediction method.

### 2.5. Transmembrane Topology and Antigenicity Prediction of the Selected Epitopes

The transmembrane topology of the selected epitopes were determined using the transmembrane topology of protein helices determinant, TMHMM v2.0 server (http://www.cbs.dtu.dk/services/TMHMM/). The server predicts whether the epitope would be transmembrane or it would remain inside or outside of the membrane [20]. The antigenicity of the selected epitopes were predicted using VaxiJen v2.0 (http://www.ddg-pharmfac.net/vaxijen/VaxiJen/VaxiJen.htm), using the tumor model and threshold of 0.4.

### 2.6. Allergenicity and Toxicity Prediction of the Epitopes

The allergenicity of the selected epitopes were predicted using two online tools, AllerTOP v2.0 (https://www.ddg-pharmfac.net/AllerTOP/) and AllergenFP v1.0 (http://ddg-pharmfac.net/AllergenFP/). However, the results predicted by AllerTOP were given priority since the server has better accuracy of 88.7% than AllergenFP server (87.9%) [21, 22]. The toxicity prediction of the selected epitopes were carried out using ToxinPred server (http://crdd.osdd.net/raghava/toxinpred/), using SVM (Swiss-Prot) based method, keeping all the parameters default.

### 2.7. Conservancy Prediction of the Selected Epitopes

The conservancy analysis of the selected epitopes were performed using the epitope conservancy analysis tool of IEDB server (https://www.iedb.org/conservancy/) (Vita et al., 2018). The sequence identity threshold was kept ‘>=50’. For the conservancy analysis of the selected epitopes of the Ebola virus strain Mayinga-76, the matrix protein VP40, envelope glycoprotein and nucleoprotein of Ebola virus strain Gabon-94 and Kikwit-95 were used for comparison along with the proteins of the Ebola virus strain Mayinga-76 itself (UniProt accession numbers: Q05128, Q2PDK5, Q77DJ6, Q05320, O11457, P87666, Q9QCE9, P18272 and O72142). Based on the antigenicity, allergenicity, toxicity and conservancy analysis, the best ligands were selected for the further analysis and vaccine construction. The T-cell epitopes that showed antigenicity, non-allergenicity, non-toxicity and high (more than 90%) conservancy and more than 50% minimum identity, were considered as the best epitopes. For B-cell epitope selection, only the antigenic and non-allergenic epitopes were taken for further analysis.

### 2.8. Cluster Analysis of the MHC Alleles

Cluster analysis of the MHC alleles helps to identify the alleles of the MHC class-I and class-II molecules that have similar binding specificities. The cluster analysis of the MHC alleles were carried out by online tool MHCcluster 2.0 (http://www.cbs.dtu.dk/services/MHCcluster/) [23]. During the analysis, the number of peptides to be included was kept 50,000, the number of bootstrap calculations were kept 100 and all the HLA supertype representatives (MHC class-I) and HLA-DR representatives (MHC class-II) were selected. For analyzing the MHC class-I alleles, the NetMHCpan-2.8 prediction method was used. The output of the server generated results in the form of MHC specificity tree and MHC specificity heat-map.

### 2.9. Generation of the 3D Structures of the Selected Epitopes

The 3D structures of the selected best epitopes were generated using online 3D generating tool PEP-FOLD3 (http://bioserv.rpbs.univ-paris-diderot.fr/services/PEP-FOLD3/). The server is a tool for generating de novo peptide 3D structure [24, 25, 26].

### 2.10. Molecular Docking of the Selected Epitopes

The molecular docking of the selected epitopes were carried out by online docking tool PatchDock (https://bioinfo3d.cs.tau.ac.il/PatchDock/php.php). PatchDock tool has algorithms that divide the Connolly dot surface representation of the molecules into concave, convex and flat patches. After that the complementary patches are matched by the server for generating potential candidate transformations. Next, each of the candidate transformations is evaluated by scoring function and finally, an RMSD (root mean square deviation) clustering is applied to the candidate solutions for discarding the redundant solutions. The top scored solutions are made the top ranked solutions by the server. After docking by PatchDock, the docking results were refined and re-scored by FireDock server (http://bioinfo3d.cs.tau.ac.il/FireDock/php.php). The FireDock server generates global energies upon the refinement of the best solutions from the PatchDock server and ranks them based on the generated global energies and the lowest global energy is always appreciable and preferred [27, 28, 29, 30]. The molecular docking experiments were carried out using the HLA-A*11-01 allele (PDB ID: 5WJL) and HLA DRB1*04-01 (PDB ID: 5JLZ) as receptors and the ligands were the best selected MHC-I and MHC-II epitopes, respectively. The receptors were downloaded from the RCSB-Protein Data Bank (https://www.rcsb.org/) server. The best results were visualized using Discovery Studio Visualizer (Dassault Systèmes BIOVIA, Discovery Studio Visualizer, v19.1.0.1828 (2019), San Diego: Dassault Systèmes).

### 2.11. Vaccine Construction

Three possible vaccines were constructed against the selected Ebola virus strain Mayinga-76. For successful vaccine construction, the predicted CTL, HTL and BCL epitopes were conjugated together. All the vaccines were constructed maintaining the sequence: adjuvant, PADRE sequence, CTL epitopes, HTL epitopes and BCL epitopes. Three different adjuvant sequences were used for vaccine construction: beta defensin, L7/L12 ribosomal protein and HABA protein (*M. tuberculosis*, accession number: AGV15514.1). By acting as agonist, beta-defensin adjuvants stimulate the activation of the toll like receptors (TLRs): 1, 2 and 4. The L7/L12 ribosomal protein and HABA protein have the ability to activate TLR-4. During the vaccine construction, various linkers were used: EAAAK linkers were used to conjugate the adjuvant and PADRE sequence, GGGS linkers were used to attach the PADRE sequence with the CTL epitopes and the CTL epitopes with the other CTL epitopes, GPGPG linkers were used to connect the CTL epitopes with the HTL epitopes and also the HTL epitopes among themselves. The KK linkers were used for conjugating the HTL and BCL epitopes as well as the BLC epitopes among themselves [31, 32, 33, 34, 35, 36, 37]. The PADRE sequence improves the CTL response of the vaccines that contain the PADRE sequence [38]. Total three vaccines were constructed in the experiment.

### 2.12. Antigenicity, Allergenicity and Physicochemical Property Analysis

The antigenicity of the constructed vaccines were determined by the online server VaxiJen v2.0 (http://www.ddg-pharmfac.net/vaxijen/VaxiJen/VaxiJen.htm). The threshold of the prediction was kept at 0.4 [15, 16, 17]. AlgPred (http://crdd.osdd.net/raghava/algpred/) and AllerTop v2.0 (https://www.ddg-pharmfac.net/AllerTOP/) were used for the prediction of the allergenicity of the vaccine constructs. The AlgPred server predicts the possible allergens based on similarity of known epitope of any of the known region of the protein [39]. MEME/MAST motif prediction approach is used in the allergenicity prediction of the vaccines by AlgPred. Moreover, various physicochemical properties of the vaccines were examined by the online server ProtParam (https://web.expasy.org/protparam/) [18].

### 2.13. Secondary and Tertiary Structure Prediction of the Vaccine Constructs

The secondary structures of the vaccine constructs were generated using online tool PRISPRED (http://bioinf.cs.ucl.ac.uk/psipred/). PRISPRED is a simple secondary structure generator that can be used to predict the transmembrane topology, transmembrane helix, fold and domain recognition etc. along with the secondary structure prediction [40, 41]. The PRISPRED 4.0 prediction method was used to predict the secondary structures of the vaccine constructs. The β-sheet structure of the vaccines were determined by another online tool, NetTurnP v1.0 (http://www.cbs.dtu.dk/services/NetTurnP/) [42]. The tertiary or 3D structures of the vaccines were generated using online tool RaptorX (http://raptorx.uchicago.edu/) server. The server is a fully annotated tool for the prediction of protein structure, the property and contact prediction, sequence alignment etc. [43, 44, 45].

### 2.14. 3D Structure Refinement and Validation

The 3D structures of the constructed vaccines were refined using online refinement tool 3Drefine (http://sysbio.rnet.missouri.edu/3Drefine/). The server is a quick, easy and efficient tool for protein structure refinement [46]. For each of the vaccine, the refined model 1 was downloaded for validation. The refined vaccine proteins were then validated by analyzing the Ramachandran plots which were generated using the online tool, PROCHECK (https://servicesn.mbi.ucla.edu/PROCHECK/) [47, 48].

### 2.15. Vaccine Protein Disulfide Engineering

The vaccine protein disulfide engineering was carried out by online tool Disulfide by Design 2 v12.2 (http://cptweb.cpt.wayne.edu/DbD2/). The server predicts the possible sites within a protein structure that may undergo disulfide bond formation [49]. When engineering the disulfide bonds, the intra-chain, inter-chain and C_β_ for glycine residue were selected. The χ_3_ angle was kept -87° or +97° ±5 and C_α_-C_β_-S_γ_ angle was kept 114.6° ±10, during the experiment.

### 2.16. Protein-Protein Docking

In protein-protein docking, the constructed Ebola virus vaccines were analyzed by docking against various MHC alleles and toll like receptors (TLR). One best vaccine would be selected based on its superior performances in the docking experiment. When viral infections occur, the viral particles or antigens are recognized by the MHC complex. The various segments of the MHC molecules are encoded by different alleles. For this reason, the vaccines should have good binding affinity with these MHC portions that are encoded by different alleles [50]. All the vaccine constructs were docked against the selected MHC alleles to test their binding affinity. In this experiment, the vaccines constructs were docked against DRB1*0101 (PDB ID: 2FSE), DRB3*0202 (PDB ID: 1A6A), DRB5*0101 (PDB ID: 1H15), DRB3*0101 (PDB ID: 2Q6W), DRB1*0401 (PDB ID: 2SEB), and DRB1*0301 (PDB ID: 3C5J). Moreover, studies have proved that TLR-8, present on the immune cells, are responsible for mediating the immune responses against the RNA viruses and TLR-3 of the immune cells mediates immune responses against the DNA viruses [51, 52]. The ebola virus is a RNA virus [53]. For this reason, the vaccine constructs of ebola virus were also docked against TLR-8 (PDB ID: 3W3M). The protein-protein docking was carried out using various online docking tools. The docking was carried out three times by three different online servers for improving the accuracy of the docking. First, the docking was carried out by ClusPro 2.0 (https://cluspro.bu.edu/login.php). The server ranks the clusters of docked complexes based on their center and lowest energy scores. However, these scores do not represent the actual binding affinity of the proteins with their targets [54, 55, 56]. The bonding affinity (ΔG in kcal mol^-1^) of docked complexes were generated by PRODIGY tool of HADDOCK webserver (https://haddock.science.uu.nl/). The lower the binding energy generated by the server, the higher the binding affinity [57, 58, 59]. Moreover, the docking was again performed by PatchDock (https://bioinfo3d.cs.tau.ac.il/PatchDock/php.php) and later refined and re-scored by FireDock server (http://bioinfo3d.cs.tau.ac.il/FireDock/php.php). The FireDock server ranks the docked complexes based on their global energy and the lower the score, the better the result [27, 28, 29, 30] Later, the docking was performed using HawkDock server (http://cadd.zju.edu.cn/hawkdock/). The Molecular Mechanics/Generalized Born Surface Area (MM-GBSA) study was also carried out using HawkDock server. According to the server, the lower scores and lower energy corresponds to better scores [60, 61, 62, 63]. The HawkDock server generates several models of docked complex and ranks them by assigning HawkDock scores in the ascending order. For each of the vaccines and their respective targets, the score of model 1 was taken for analysis. Furthermore, the model 1 of every complex was analyzed for MM-GBSA study. From the docking experiment, one best vaccine was selected for further analysis. The docked structures were visualized by PyMol tool [64].

### 2.17. Molecular Dynamic Simulation

The molecular dynamics simulation study was conducted for the best selected vaccine, by the online server iMODS (http://imods.chaconlab.org/). iMODS server is a fast, online, user-friendly and effective molecular dynamics simulation server that can be used efficiently to investigate the structural dynamics of the protein complexes. The server provides the values of deformability, B-factor (mobility profiles), eigenvalues, variance, co-variance map and elastic network. For a protein complex, the deformability depends on the ability to deform at each of its amino acid. The eigenvalue is related with the energy that is required to deform the given structure and the lower the eigenvalue, the easier the deformability of the complex. The eigenvalue also represents the motion stiffness of the protein complex. The server is a fast and easy tool for determining and measuring the protein flexibility [65, 66, 67, 68, 69]. For analysing the molecular dynamics simulation of the EV-1-TLR-8 docked complex was used. The docked PDB files were uploaded to the iMODS server and the results were displayed keeping all the parameters as default.

### 2.18. Codon Adaptation and In Silico Cloning

In codon adaptation, the best selected vaccine protein was reverse transcribed to the possible DNA sequence. The DNA sequence should encode the target vaccine protein. After that, the reverse transcribed DNA sequence was adapted according to the desired organism, so that the cellular mechanisms of that specific organism could use the codons of the adapted DNA sequences efficiently and provide better production of the desired product. Codon adaptation is a necessary step of in silico cloning because the same amino acid can be encoded by different codons in different organisms (codon biasness). Moreover, the cellular mechanisms of an organism may be different from another organism and a codon for a specific amino acid may not work in another organism. For this reason, codon adaptation can predict the suitable codon that can encode a specific amino acid in a specific organism [70, 71]. The predicted protein sequences of the best selected vaccines were used for codon adaptation by the Java Codon Adaptation Tool or JCat server (http://www.jcat.de/) [70]. Eukaryotic *E. coli* strain K12 was selected and rho-independent transcription terminators, prokaryotic ribosome binding sites and SgrA1 and SphI cleavage sites of restriction enzymes, were avoided. In the JCat server, the protein sequences were reverse translated to the optimized possible DNA sequences. The optimized DNA sequences were taken and SgrA1 and SphI restriction sites were conjugated at the N-terminal and C-terminal sites, respectively. Next, the SnapGene [72] restriction cloning module was used to insert the new adapted DNA sequences between SgrA1 and SphI of pET-19b vector.

## 3. Results

### 3.1. Identification, Selection and Retrieval of Viral Protein Sequences

The Zaire Ebola virus, strain Mayinga-76 was identified selected from the NCBI (https://www.ncbi.nlm.nih.gov/). Three proteins from the viral structures were selected for the possible vaccine construction. These proteins were: envelope glycoprotein (accession number: Q05320), matrix protein VP40 (accession number: Q05128) and nucleoprotein (accession number: P18272). The protein sequences were retrieved from the online server, UniProt (https://www.uniprot.org/). The protein sequences in fasta format:

>sp|Q05128|VP40_EBOZM Matrix protein VP40 OS=Zaire ebolavirus (strain Mayinga-76) OX=128952 GN=VP40 PE=1 SV=1 MRRVILPTAPPEYMEAIYPVRSNSTIARGGNSNTGFLTPESVNGDTPSNPLRPIADDTIDHASHTPGSVSSAFILEAMVNVISGPKVLMKQIPIWLPLGVADQKTYSFDSTTAAIMLASYTITHFGKATNPLVRVNRLGPGIPDHPLRLLRIGNQAFLQEFVLPPVQLPQYFTFDLTALKLITQPLPAATWTDDTPTGSNGALRPGISFHPKLRPILLPNKSGKKGNSADLTSPEKIQAIMTSLQDFKIVPIDPTKNIMGIEVPETLVHKLTGKKVTSKNGQPIIPVLLPKYIGLDPVAPGDLTMVITQDCDTCHSPASLPAVIEK

>sp|Q05320|VGP_EBOZM Envelope glycoprotein OS=Zaire ebola virus (strain Mayinga-76) OX=128952 GN=GP PE=1 SV=1 MGVTGILQLPRDRFKRTSFFLWVIILFQRTFSIPLGVIHNSTLQVSDVDKLVCRDKLSSTNQLRSVGLNLEGNGVATDVPSATKRWGFRSGVPPKVVNYEAGEWAENCYNLEIKKPDGSECLPAAPDGIRGFPRCRYVHKVSGTGPCAGDFAFHKEGAFFLYDRLASTVIYRGTTFAEGVVAFLILPQAKKDFFSSHPLREPVNATEDPSSGYYSTTIRYQATGFGTNETEYLFEVDN LTYVQLESRFTPQFLLQLNETIYTSGKRSNTTGKLIWKVNPEIDTTIGEWAFWETKKNLTRKIRSEELSFTVVSNGAKNISGQSPARTSSDPGTNTTTEDHKIMASENSSAMVQVHSQGREAAVSHLTTLATISTSPQSLTTKPGPDNSTHNTPVYKLDISEATQVEQHHRRTDNDSTASDTPSATTAAGPPKAENTNTSKSTDFLDPATTTSPQNHSETAGNNNTHHQDTGEESASSGKLGLITNTIAGVAGLITGGRRTRREAIVNAQPKCNPNLHYWTTQDEGAAIGLAWIPYFGPAAEGIYIEGLMHNQDGLICGLRQLANETTQALQLFLRATTELRTFSILNRKAIDFLLQRWGGTCHILGPDCCIEPHDWTKNITDKIDQIIHDFVDKTLPDQGDNDNWWTGWRQWIPAGIGVTGVIIAVIALFCICKFVF

>sp|P18272|NCAP_EBOZM Nucleoprotein OS=Zaire ebola virus (strain Mayinga-76) OX=128952 GN=NP PE=1 SV=2 MDSRPQKIWMAPSLTESDMDYHKILTAGLSVQQGIVRQRVIPVYQVNNLEEICQLIIQAFEAGVDFQESADSFLLMLCLHHAYQGDYKLFLESGAVKYLEGHGFRFEVKKRDGVKRLEELLPAVSSGKNIKRTLAAMPEEETTEANAGQFLSFASLFLPKLVVGEKACLEKVQRQIQVHAEQGLIQYPTAWQSVGHMMVIFRLMRTNFLIKFLLIHQGMHMVAGHDANDAVISNSVAQARFSGLLIVKTVLDHILQKTERGVRLHPLARTAKVKNEVNSFKAALSSLAKHGEYAPFARLLNLSGVNNLEHGLFPQLSAIALGVATAHGSTLAGVNVGEQYQQLREAATEAEKQLQQYAESRELDHLGLDDQEKKILMNFHQKKNEISFQQTNAMVTLRKERLAKLTEAITAASLPKTSGHYDDDDDIPFPGPINDDDNPGHQDDDPTDSQDTTIPDVVVDPDDGSYGEYQSYSENGMNAPDDLVLFDLDEDDEDTKPVPNRSTKGGQQKNSQKGQHIEGRQTQSRPIQNVPGPHRTIHHASAPLTDNDRRNEPSGSTSPRMLTPINEEADPLDDADDETSSLPPLESDDEEQDRDGTSNRTPTVAPPAPVYRDHSEKKELPQDEQQDQDHTQEARNQDSDNTQSEHSF EEMYRHILRSQGPFDAVLYYHMMKDEPVVFSTSDGKEYTYPDSLEEEYPPWLTEKEAMNEENRFVTLDGQQFYWPVMNHKNKFMAILQHHQ

### 3.2. Antigenicity Prediction and Physicochemical Property Analysis of the Protein Sequences

VaxiJen v2.0 (http://www.ddg-pharmfac.net/vaxijen/VaxiJen/VaxiJen.htm) and ProtParam (https://web.expasy.org/protparam/) servers were used for antigenicity analysis and physicochemical property prediction of the selected protein sequences (**Table 01**). In the physicochemical property analysis, the number of amino acids, the molecular weights, theoretical pI, extinction coefficients, half-lives, instability indexes, aliphatic indexes and grand average of hydropathicity (GRAVY) of the three proteins were predicted. All the three selected protein sequences showed good level of antigenicity. The matrix protein VP40 had the lowest molecular weight of 35182.83 and only envelope glycoprotein was stable according to the prediction. All of the three proteins had 30 hours of half-life in mammalian cells. The matrix protein VP40 also showed the highest theoretical pI, aliphatic index and GRAVY values (**Table 01**).

**Table 01.**
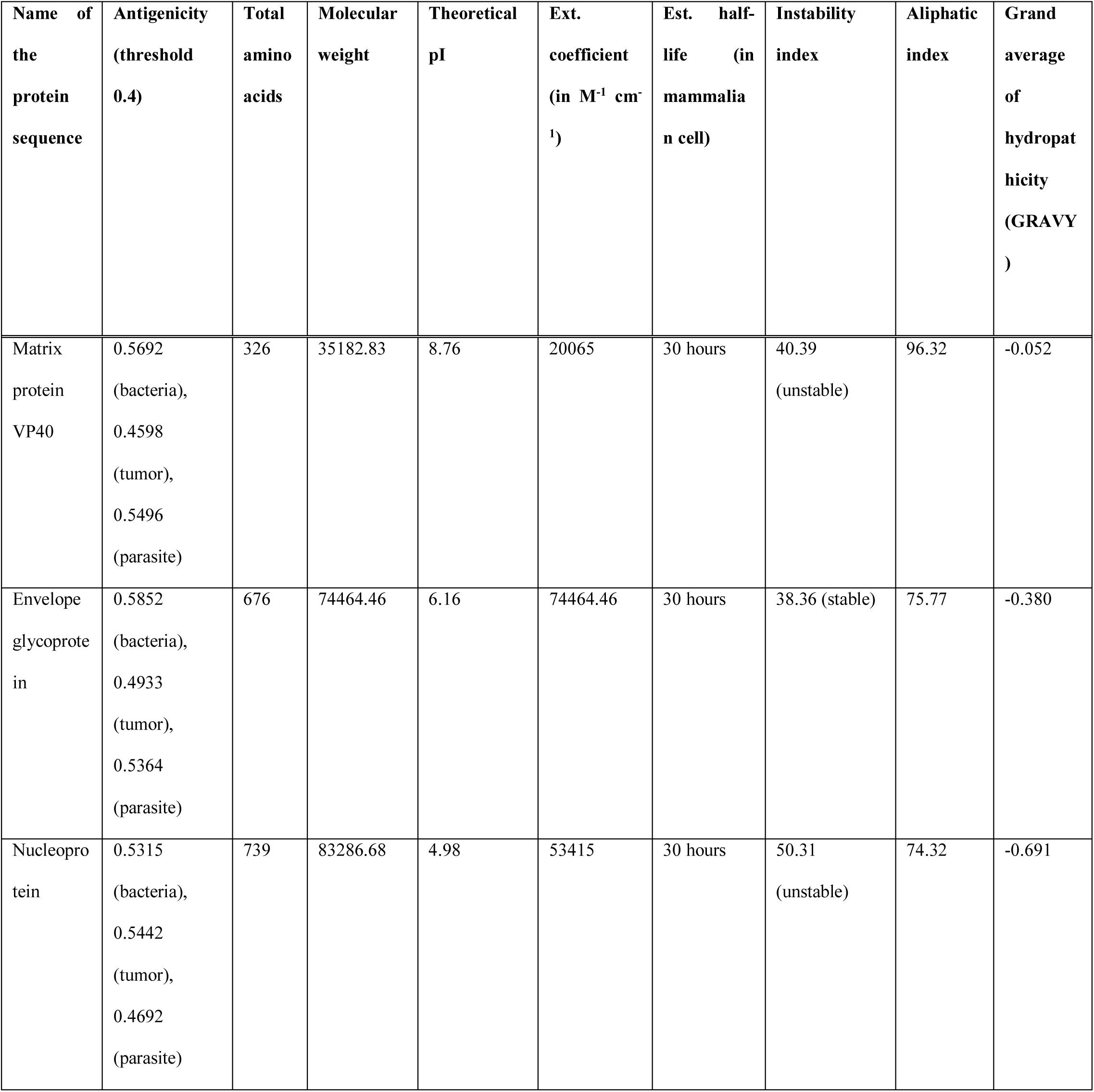
The antigenicity and physicochemical property analysis of the selected viral proteins.

### 3.3. T-cell and B-cell Epitope Prediction and Topology Determination of the Epitopes

The T-cell epitopes of MHC class-I of the three proteins were determined by NetMHCpan EL 4.0 prediction method of the IEDB (https://www.iedb.org/) server keeping the sequence length 9. The server generated over 100 such epitopes. However, based on analyzing the antigenicity scores (AS) and percentile scores, for each epitope, ten potential epitopes from the top twenty epitopes were selected randomly for antigenicity, allergenicity, toxicity and conservancy tests. The server ranks the predicted epitopes based on the ascending order of percentile scores. The lower the percentile, the better the binding affinity. The T-cell epitopes of MHC class-II (HLA DRB1*04-01 allele) of the proteins were also determined by IEDB (https://www.iedb.org/) server, where the Sturniolo prediction methods was used. For each protein, ten of the top twenty epitopes were selected randomly for further analysis. Moreover, the B-cell epitopes of the proteins were selected using Bipipered linear epitope prediction method of the IEDB (https://www.iedb.org/) server and epitopes were selected based on their higher lengths (**Figure 02**). The topology of the selected epitopes were determined by TMHMM v2.0 server (http://www.cbs.dtu.dk/services/TMHMM/). **Table 02** and **Table 03** list the potential T-cell epitopes of matrix protein VP40, **Table 04** and **Table 05** list the potential T-cell epitopes of envelope glycoprotein, **Table 06** and **Table 07** list the potential T-cell epitopes of nucleoprotein and **Table 08** list the potential B-cell epitopes with their respective topologies.

**Table 02.**
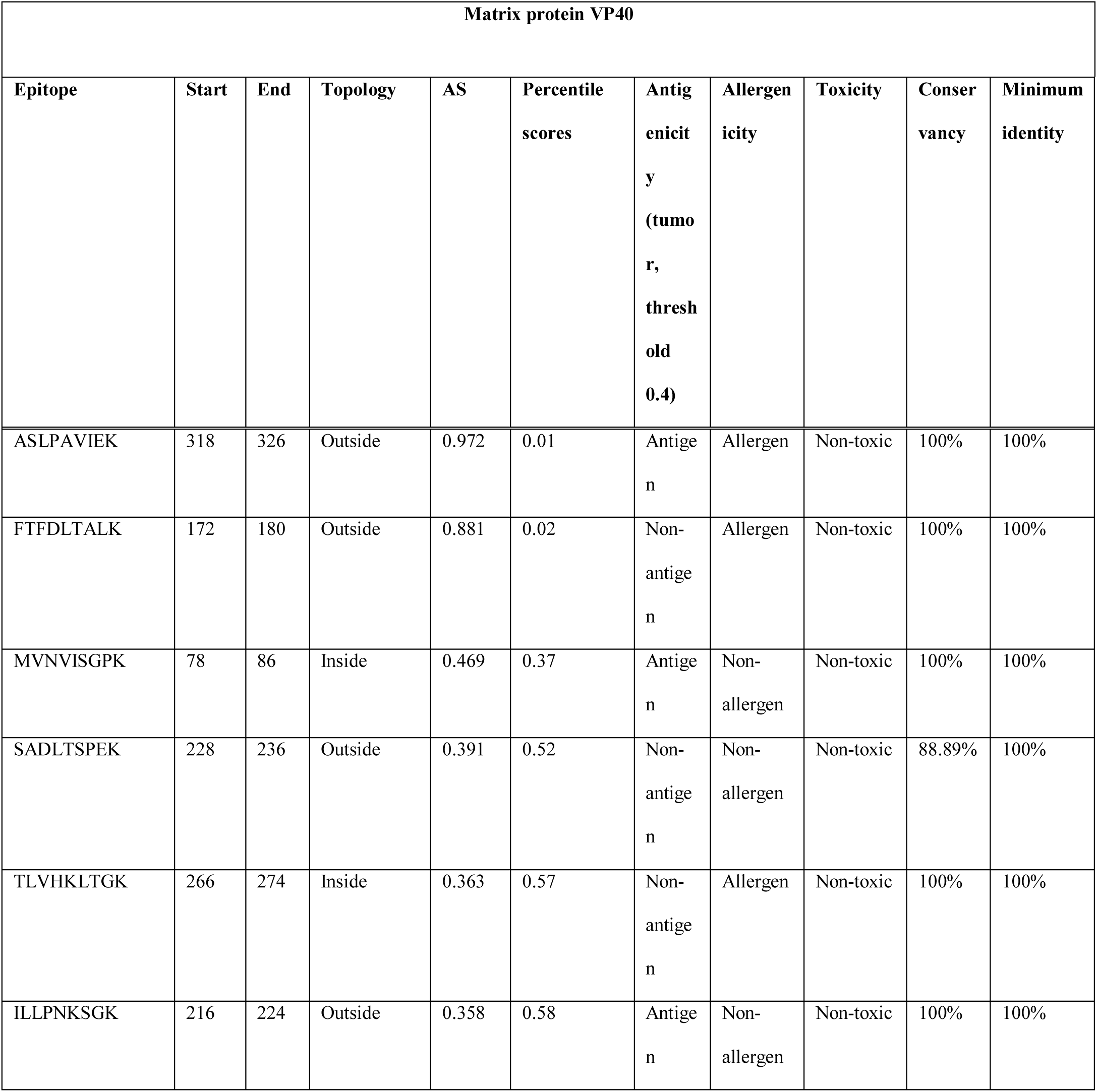
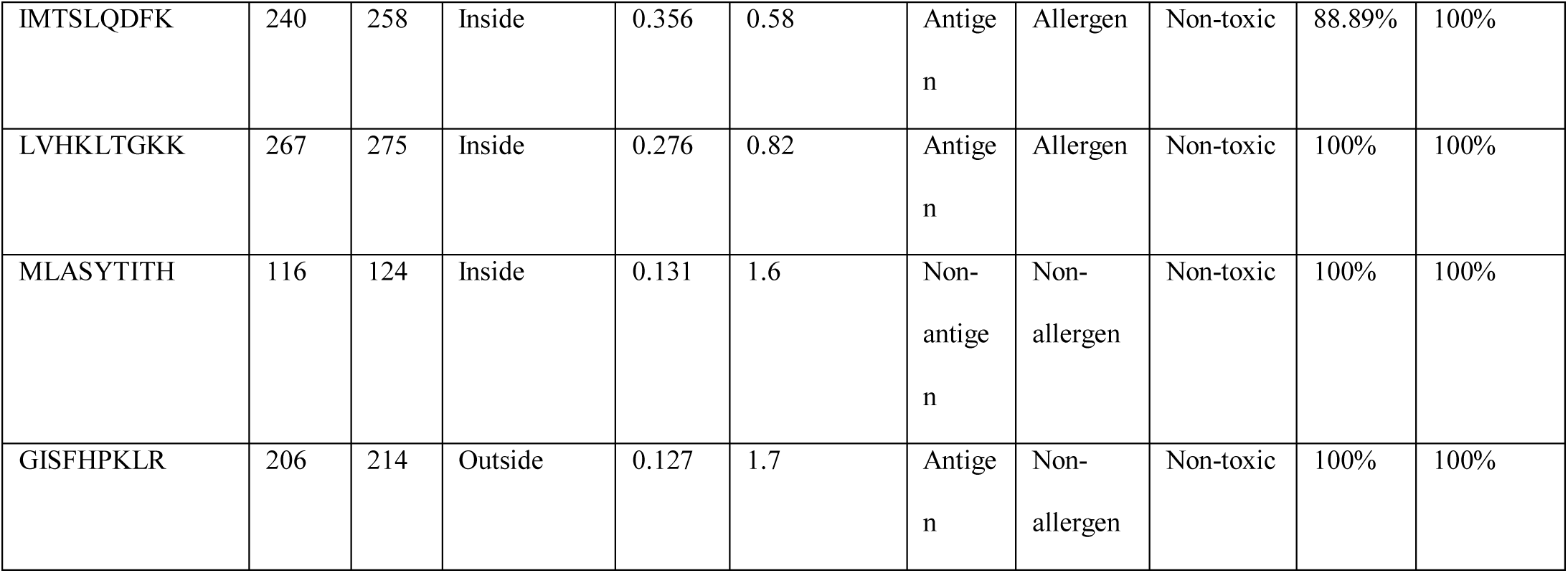
MHC class-I epitope prediction and topology, antigenicity, allergenicity, toxicity and conservancy analysis of the epitopes of matrix protein VP40.

**Table 03.**
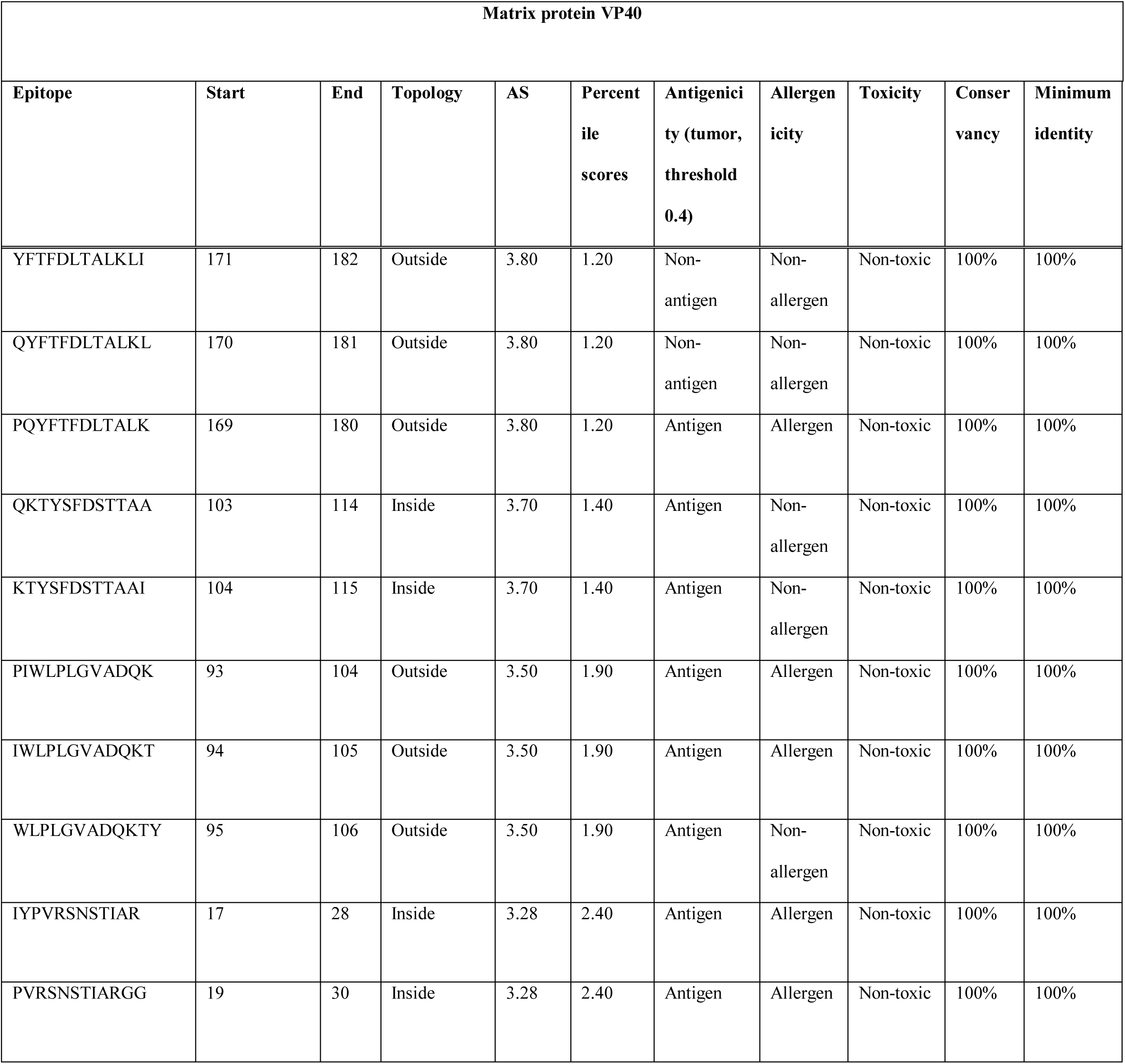
MHC class-II epitope prediction and topology, antigenicity, allergenicity, toxicity and conservancy analysis of the epitopes of matrix protein VP40.

**Table 04.**
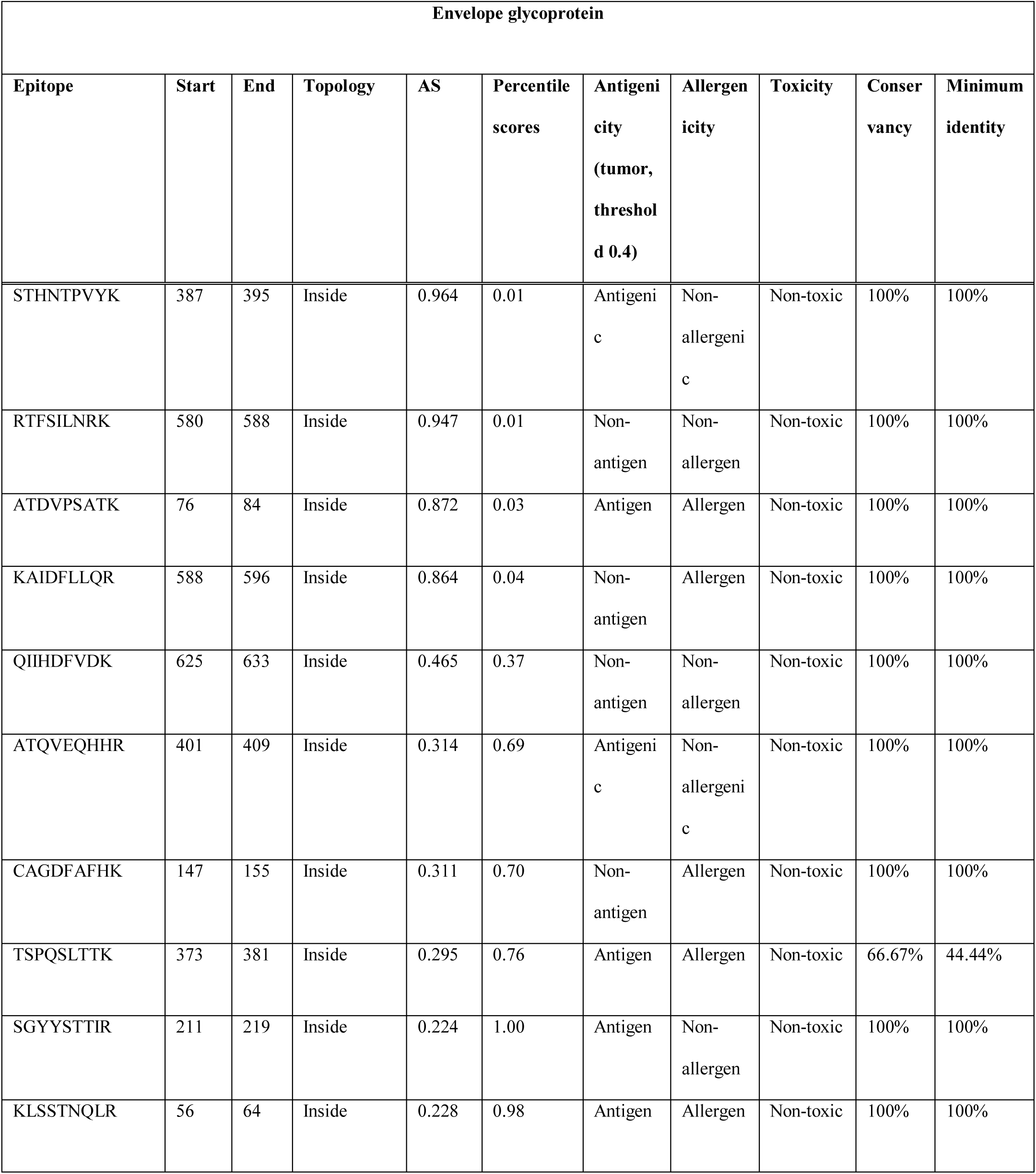
MHC class-I epitope prediction and topology, antigenicity, allergenicity, toxicity and conservancy analysis of the epitopes of envelope glycoprotein.

**Table 05.**
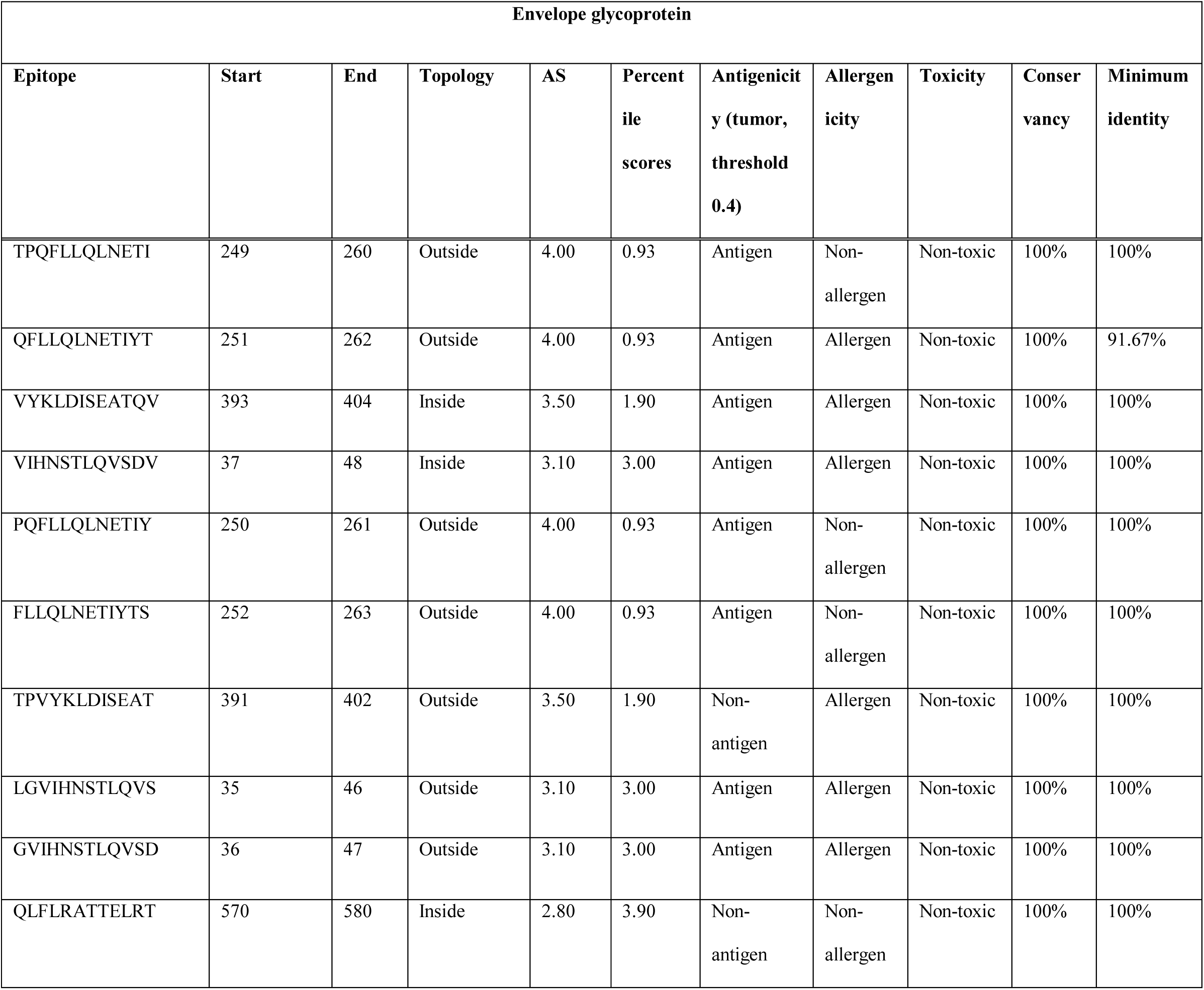
MHC class-II epitope prediction and topology, antigenicity, allergenicity, toxicity and conservancy analysis of the epitopes of envelope glycoprotein.

**Table 06.**
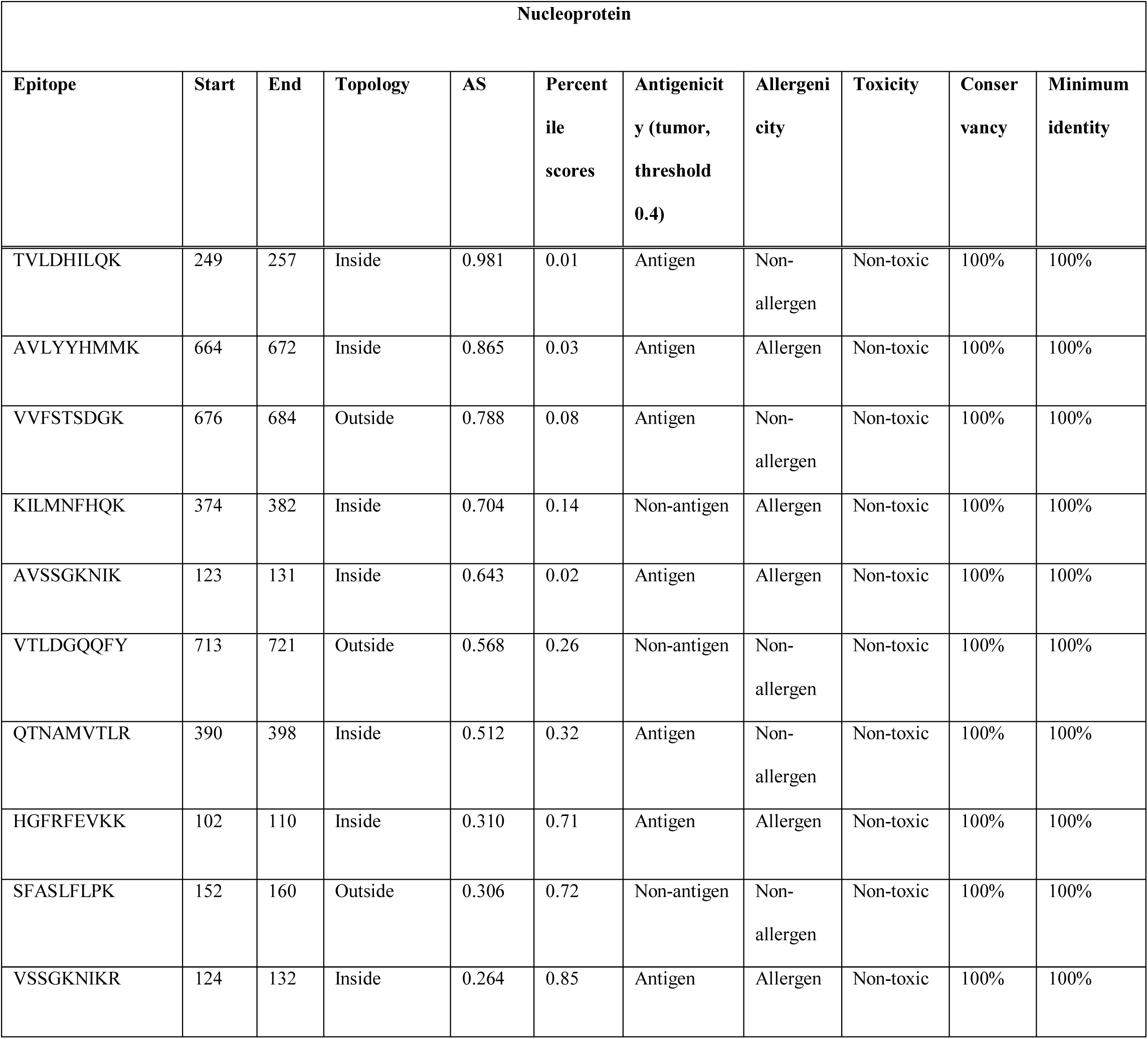
MHC class-I epitope prediction and topology, antigenicity, allergenicity, toxicity and conservancy analysis of the epitopes of nucleoprotein.

**Table 07.**
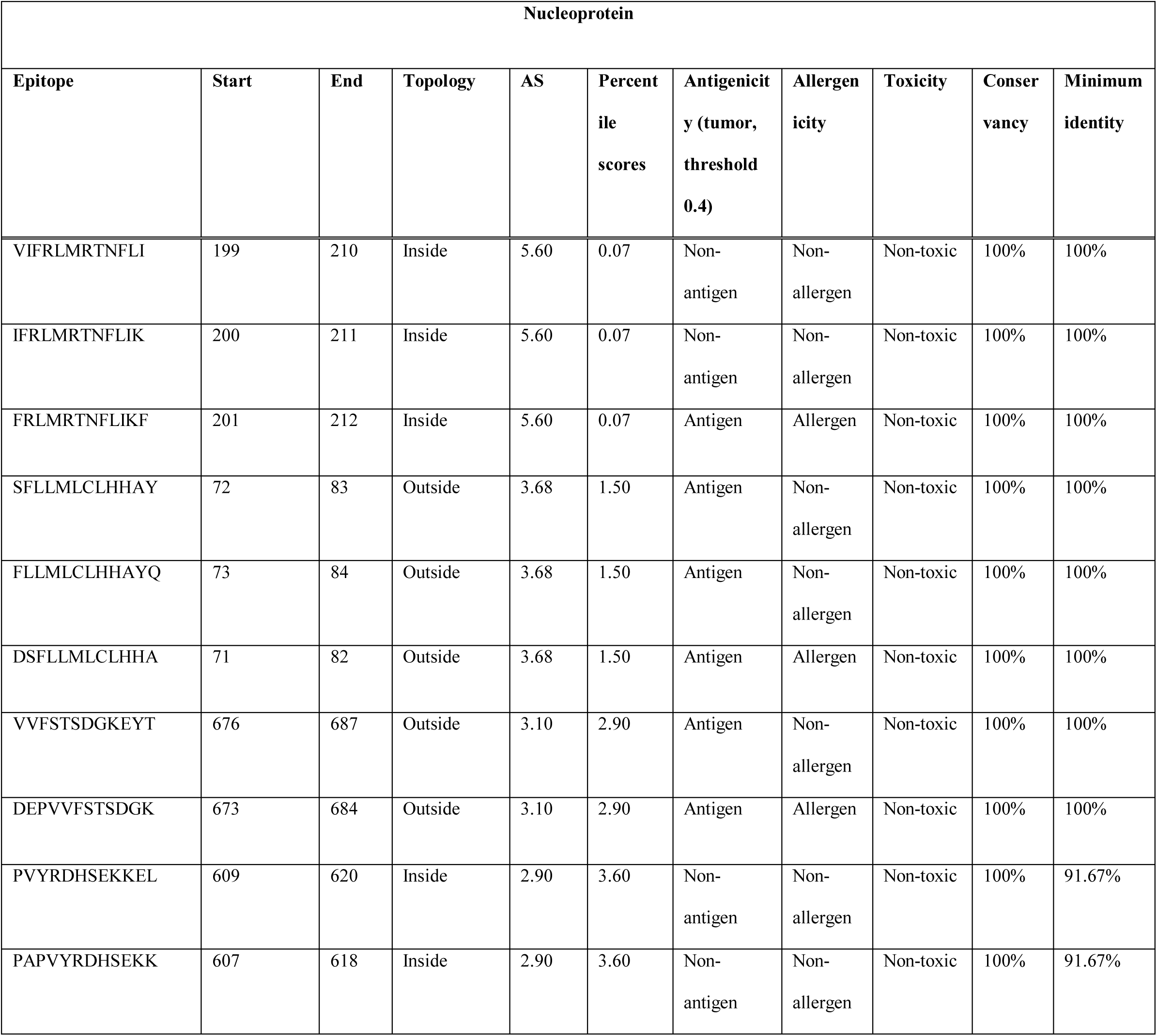
MHC class-II epitope prediction and topology, antigenicity, allergenicity, toxicity and conservancy analysis of the epitopes of nucleoprotein.

**Table 08.**
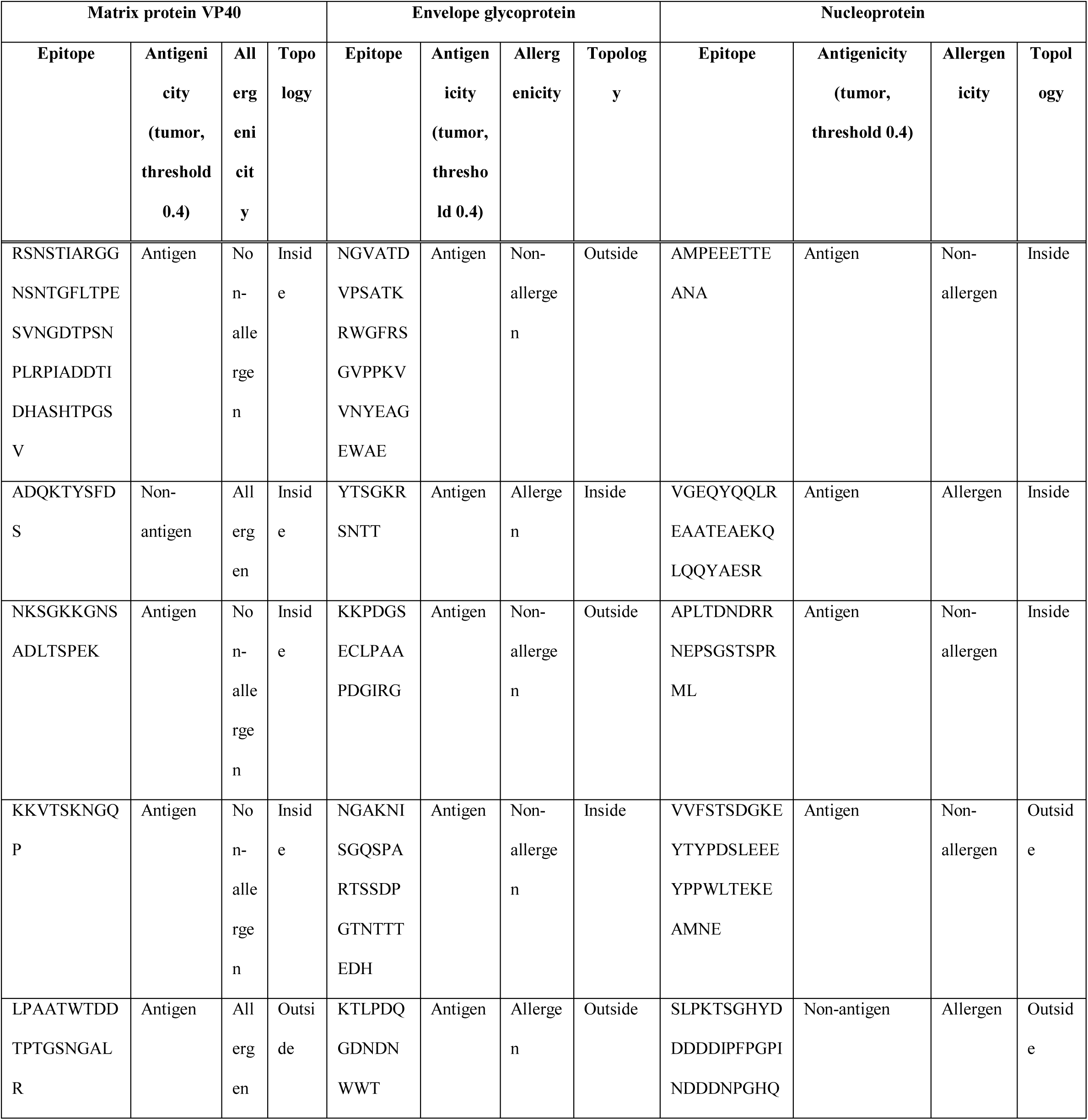
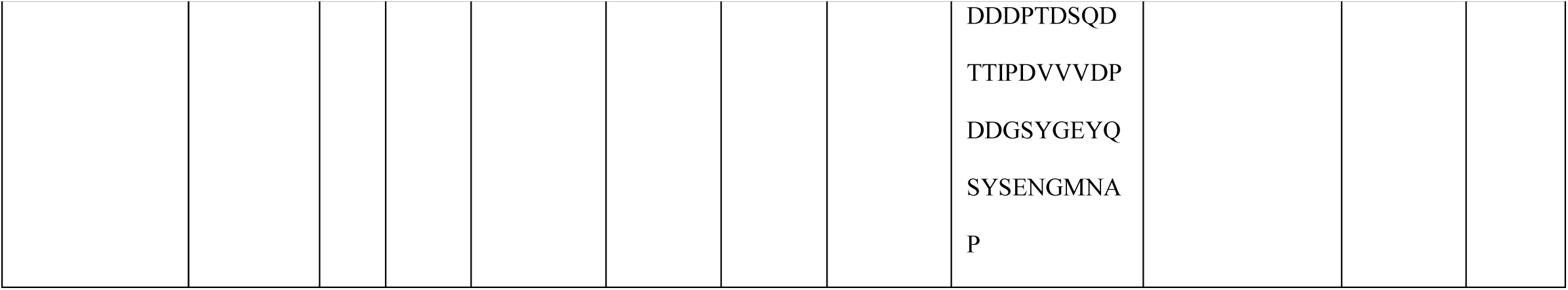
B-cell epitope prediction and antigenicity, allergenicity and topology prediction of the epitopes of the three selected proteins.

**Figure 02.**
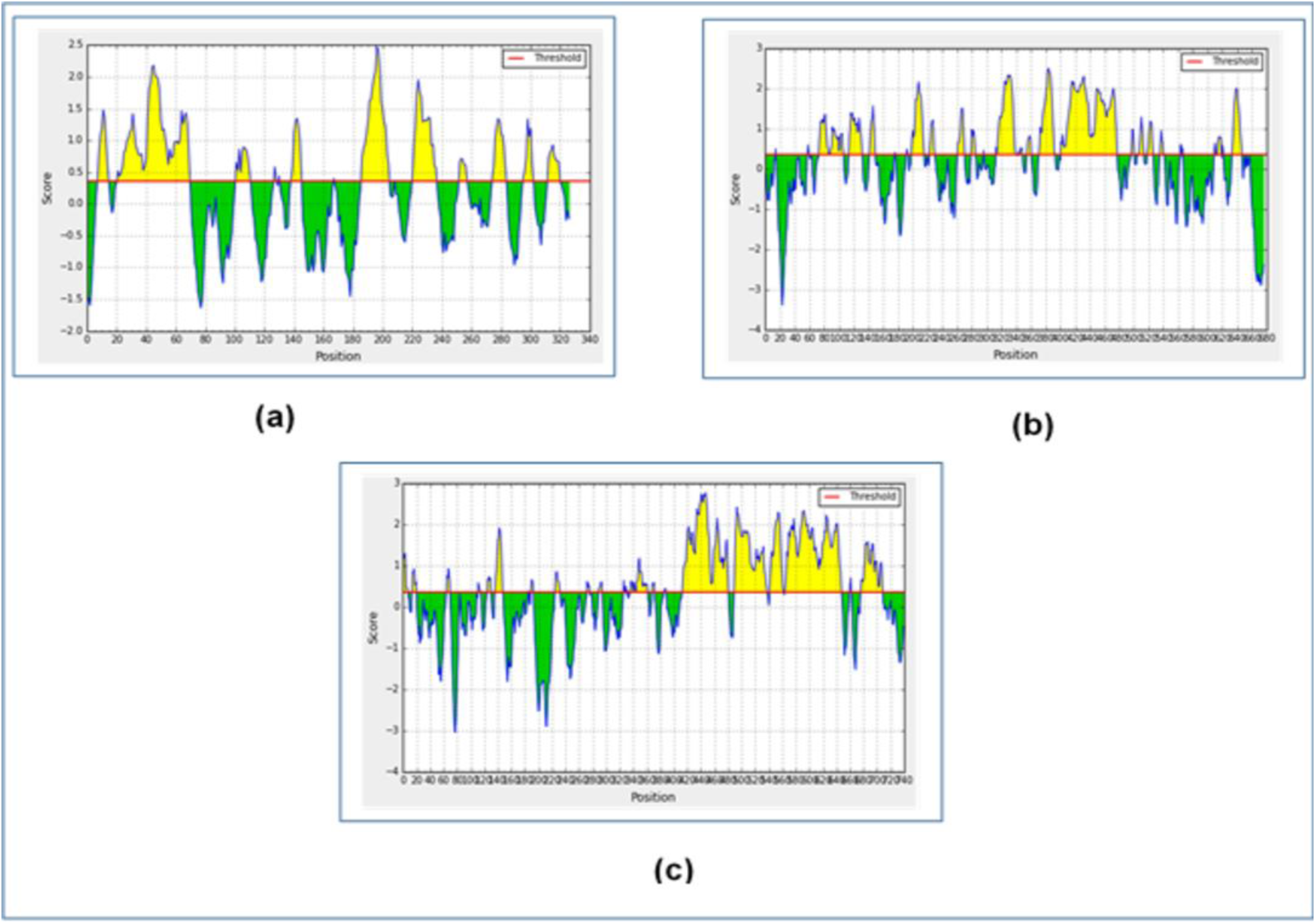
Figure showing the graphs of the B-cell epitope prediction of the three selected proteins of Ebola virus strain Mayinga-76, using Bipipered linear epitope prediction method. Here, (a) is the graph of epitope prediction for matrix protein VP40, (b) is the graph of epitope prediction for envelope glycoprotein and (c) is the graph of epitope prediction for nucleoprotein.

### 3.4. Antigenicity, allergenicity, toxicity and conservancy analysis

In the antigenicity, allergenicity, toxicity and conservancy analysis, the T-cell epitopes that were found to be highly antigenic as well as non-allergenic, non-toxic, had minimum identity of over 50% and had conservancy of over 90% as well as antigenic and non-allergenic B-cell epitopes were selected for further analysis and vaccine construction. Among the ten selected MHC class-I epitopes and ten selected MHC class-II epitopes of matrix protein VP40, total six epitopes (three epitopes from each of the category) were selected based on the mentioned criteria: MVNVISGPK, ILLPNKSGK, GISFHPKLR, QKTYSFDSTTAA, KTYSFDSTTAAI and WLPLGVADQKTY. On the other hand, among the ten selected MHC class-I epitopes and ten selected MHC class-II epitopes of envelope glycoprotein, total six epitopes (three epitopes from each of the category) were selected based on the mentioned criteria: STHNTPVYK, ATQVEQHHR, SGYYSTTIR, TPQFLLQLNETI, PQFLLQLNETIY and FLLQLNETIYTS. Moreover, like these proteins, six epitopes that obeyed the mentioned criteria, were selected for further analysis from the nucleoprotein epitopes: TVLDHILQK, VVFSTSDGK, QTNAMVTLR, SFLLMLCLHHAY, FLLMLCLHHAYQ and VVFSTSDGKEYT. For the selection of the B-cell epitopes, the highly antigenic and non-allergenic sequences were taken for vaccine construction. Three epitopes from each of the protein category were selected. For this reason, total nine epitopes B-cell epitopes were selected for vaccine construction, since they obeyed the selection criteria. (**Table 02, Table 03, Table 04, Table 05, Table 06, Table 07** and **Table 08**).

### 3.5. Cluster Analysis of the MHC Alleles

The cluster analysis of the possible MHC class-I and MHC class-II alleles that may interact with the predicted epitopes were performed by online tool MHCcluster 2.0 (http://www.cbs.dtu.dk/services/MHCcluster/). The tool generates the clusters of the alleles in phylogenetic manner. **Figure 03** illustrates the outcome of the experiment where the red zone indicates strong interaction and the yellow zone corresponds to weaker interaction.

**Figure 03.**
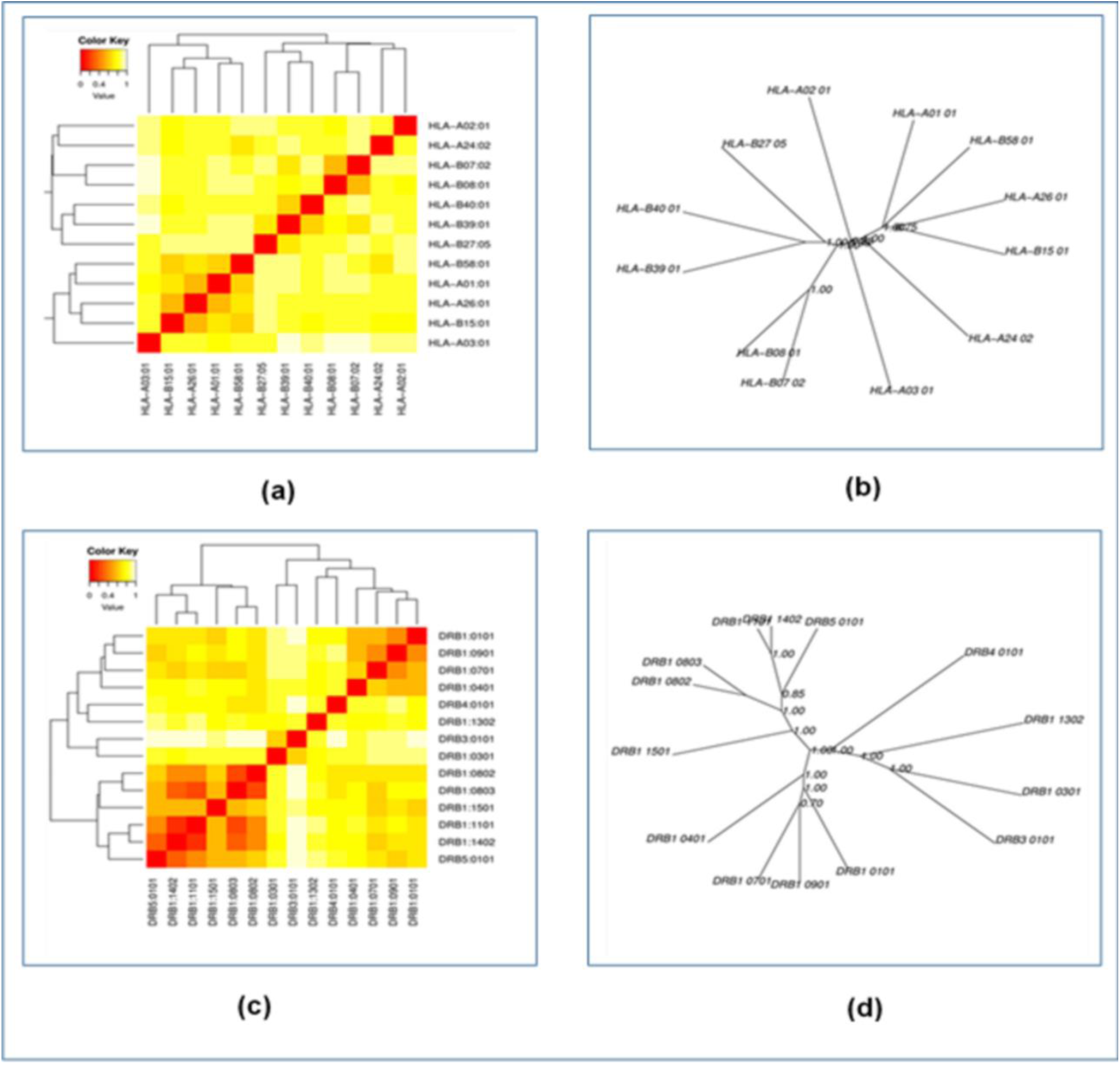
The results of the MHC cluster analysis. Here, (a) is the heat map of MHC class-I cluster analysis, (b) is the tree map of MHC class-I cluster analysis, (c) is the heat map of MHC class-II cluster analysis, (d) is the tree map of MHC class-II cluster analysis. The cluster analysis was carried out using onine server MHCcluster 2.0 (http://www.cbs.dtu.dk/services/MHCcluster/).

### 3.6. Generation of the 3D Structures of the Epitopes and Peptide-Protein Docking

All the T-cell epitopes were subjected to 3D structure generation by the PEP-FOLD3 server. The 3D structures were generated for peptide-protein docking. The docking was performed to find out, whether all the epitopes had the ability to bind with the MHC class-I and MHC class-II molecule. The selected epitopes were docked against the HLA-A*11-01 allele (PDB ID: 5WJL) and HLA DRB1*04-01 (PDB ID: 5JLZ). The docking was performed using PatchDock online docking tool and then the results were refined by FireDock online server. Among the MHC class-I epitopes of matrix protein VP40, GISFHPKLR showed the best result with the lowest global energy of -31.68. Among the MHC class-I epitopes of envelope glycoprotein, SGYYSTTIR generated the lowest and best global energy score of -42.87. QTNAMVTLR generated the best global energy score of -45.40 of the MHC class-I epitopes of nucleoprotein. Among the MHC class-II epitopes of matrix protein VP40, KTYSFDSTTAAI generated the best global energy score of -24.32 and TPQFLLQLNETI generated the lowest global energy of -2.80 and VVFSTSDGKEYT generated the lowest global energy of -12.06, among the MHC class-II epitopes of envelope glycoprotein and nucleoprotein, respectively (**Table 09**). **Figure 04** illustrates the interaction between the best docked epitopes with their respective targets.

**Table 09.**
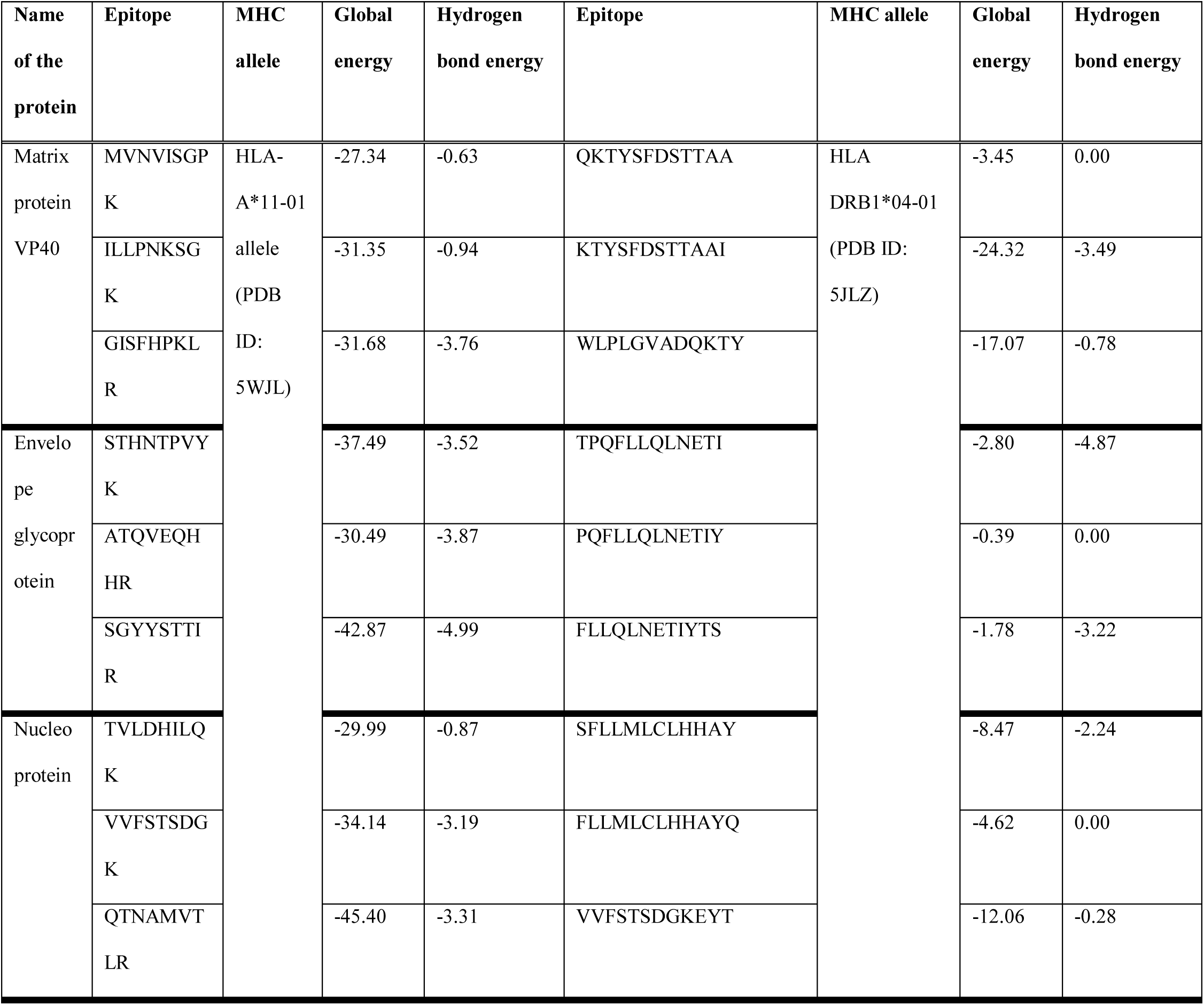
Results of molecular docking analysis of the selected epitopes.

**Figure 04.**
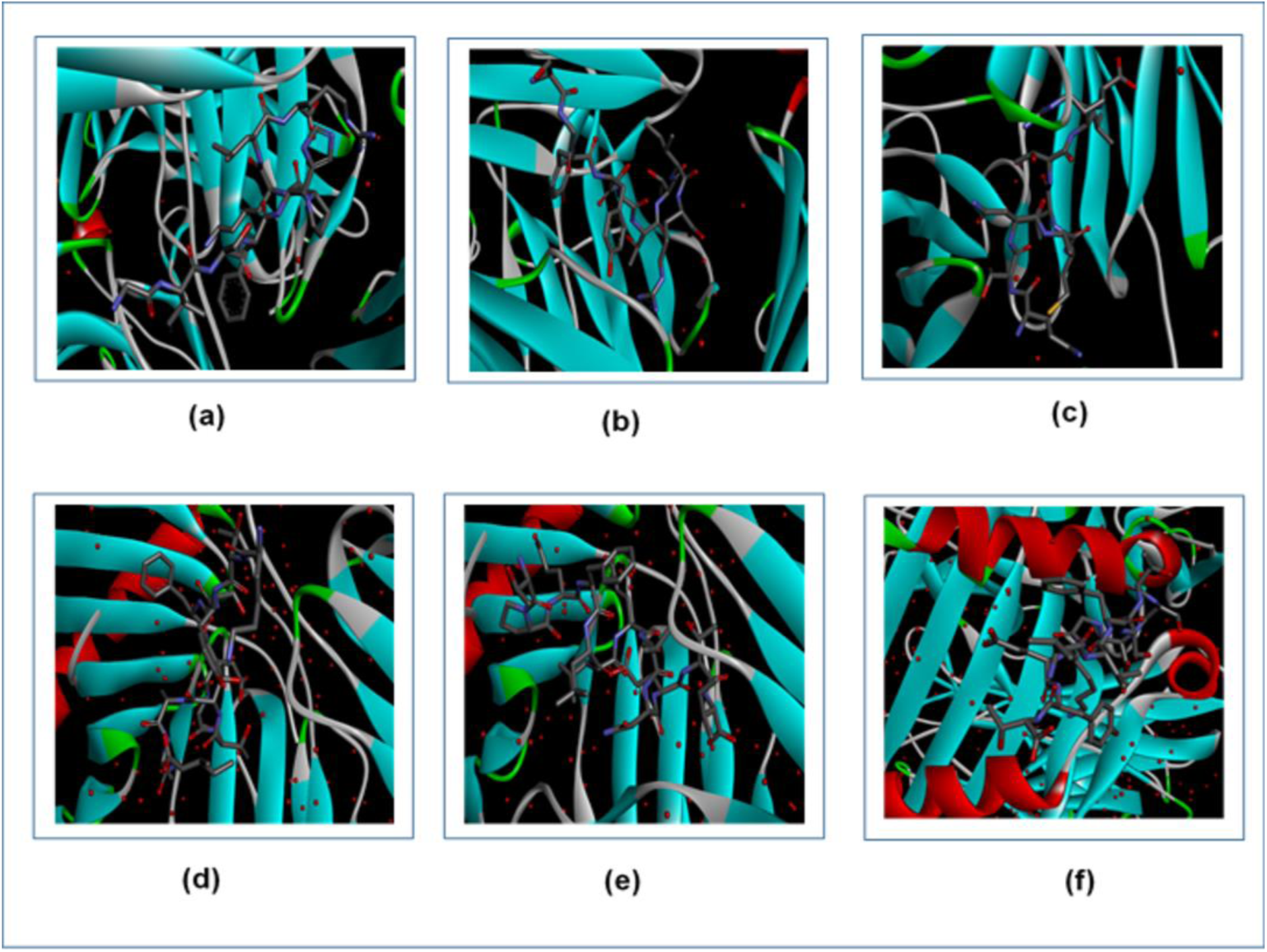
Figure showing the interactions between the best epitopes from the three proteins and their respective receptors. Here, (a) is the interaction between GISFHPKLR and MHC class-I, (b) is the interaction between SGYYSTTIR and MHC class-I, (c) is the interaction between QTNAMVTLR and MHC class-I molecule, (d) is the interaction between KTYSFDSTTAAI and MHC class-II, (e) is the interaction between TPQFLLQLNETI and MHC class-II, (f) is the interaction between VVFSTSDGKEYT and MHC class-II molecule. The interactions were visualized by Discovery Studio Visualizer.

### 3.7. Vaccine Construction

After successful docking, three vaccines were constructed, that could be used effectively to fight against ebola virus strain Mayinga-76. For vaccine constructions, three different adjuvants were used to construct three different vaccine for each of the viruses: beta defensin, L7/L12 ribosomal protein and HABA protein, were used. PADRE sequence was also used for vaccine construction. Three different vaccine constructs differed from each other only in their adjuvant sequences. To construct a vaccine, first the adjuvant sequence was conjugated with the PADRE sequence by EAAAK linker, then the PADRE sequence was added to the CTL epitopes by GGGS linkers, the CTL epitopes were also conjugated with each other by the GGGS linkers. Next, the HTL epitopes were conjugated by GPGPG linkers and the BCL epitopes were linked by KK linkers. Each vaccine construct was ended by an additional GGGS linker. The newly constructed vaccines were designated as: EV-1, EV-2 and EV-3 (**Table 10**).

**Table 10.**
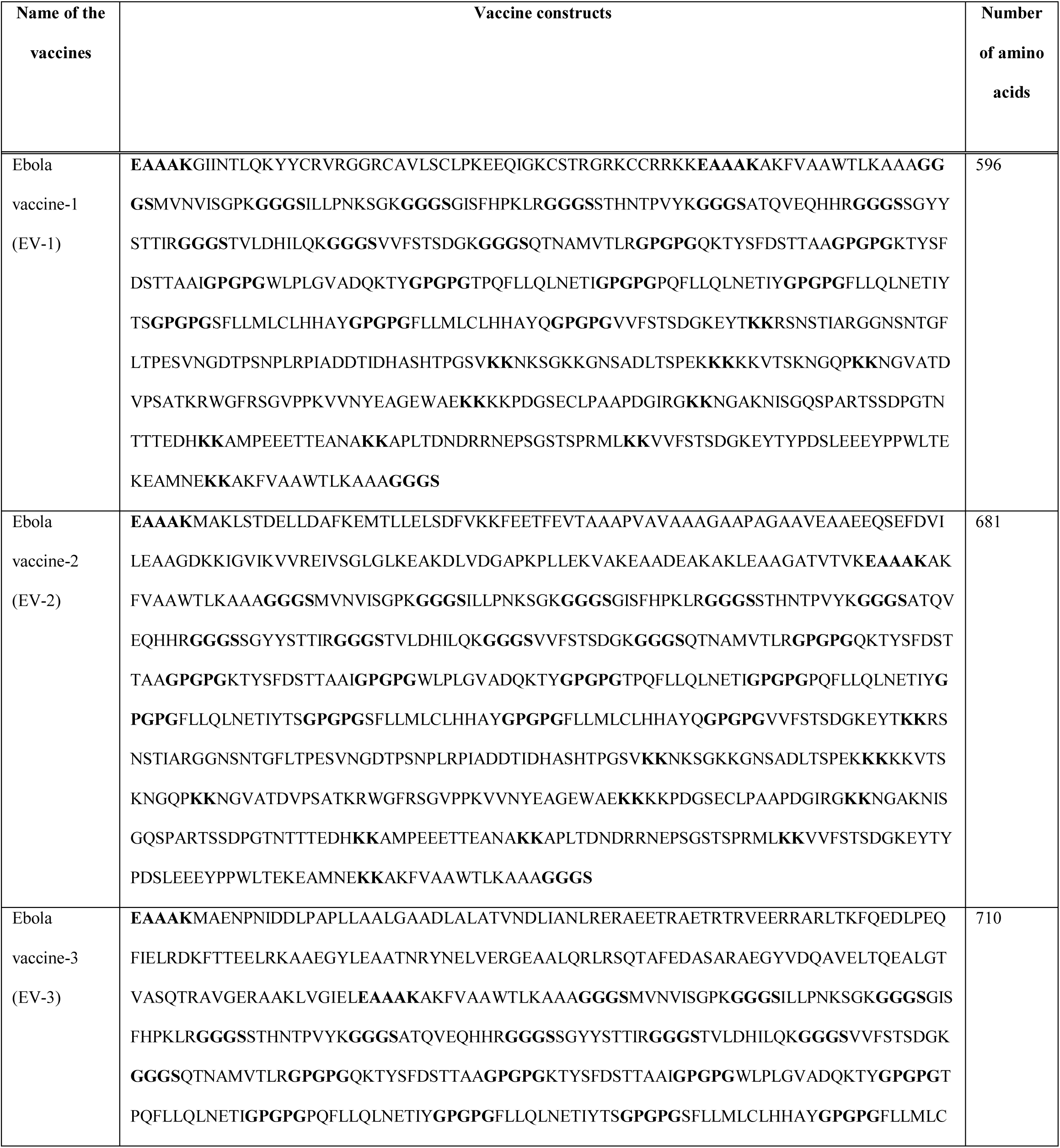
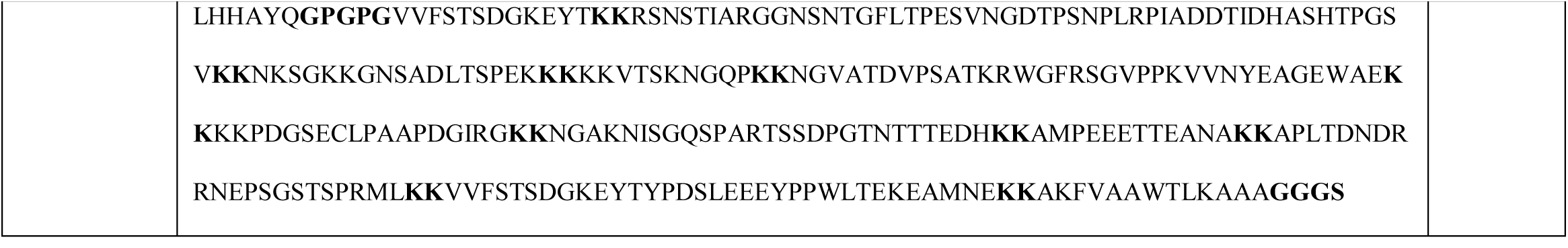
List of the vaccines constructed for ebola virus strain Mayinga-76.

### 3.8. Antigenicity, Allergenicity and Physicochemical Property Analysis of the Vaccine Constructs

The antigenicity prediction of the three vaccine constructs showed that all of the vaccine constructs were potential antigens. However, since all of the vaccine constructs were non-allergen, they are safe to use. In the physicochemical property analysis, the number of amino acids, molecular weight, extinction coefficient (in M^-1^ cm^-1^), theoretical pI, half-life, instability index, aliphatic index and GRAVY were determined. All the vaccines quite similar theoretical pI and ext. coefficient (EV-3 had the lowest value of 55475 M^-1^ cm^-1^) and all of them were stable. All of the vaccine constructs had the similar half-life of 1 hour in the mammalian cells. EV-2 had the highest GRAVY value of -0.504. The antigenicity, allergenicity and physicochemical property analysis of the three vaccine constructs are listed in **Table 11**.

**Table 11.**
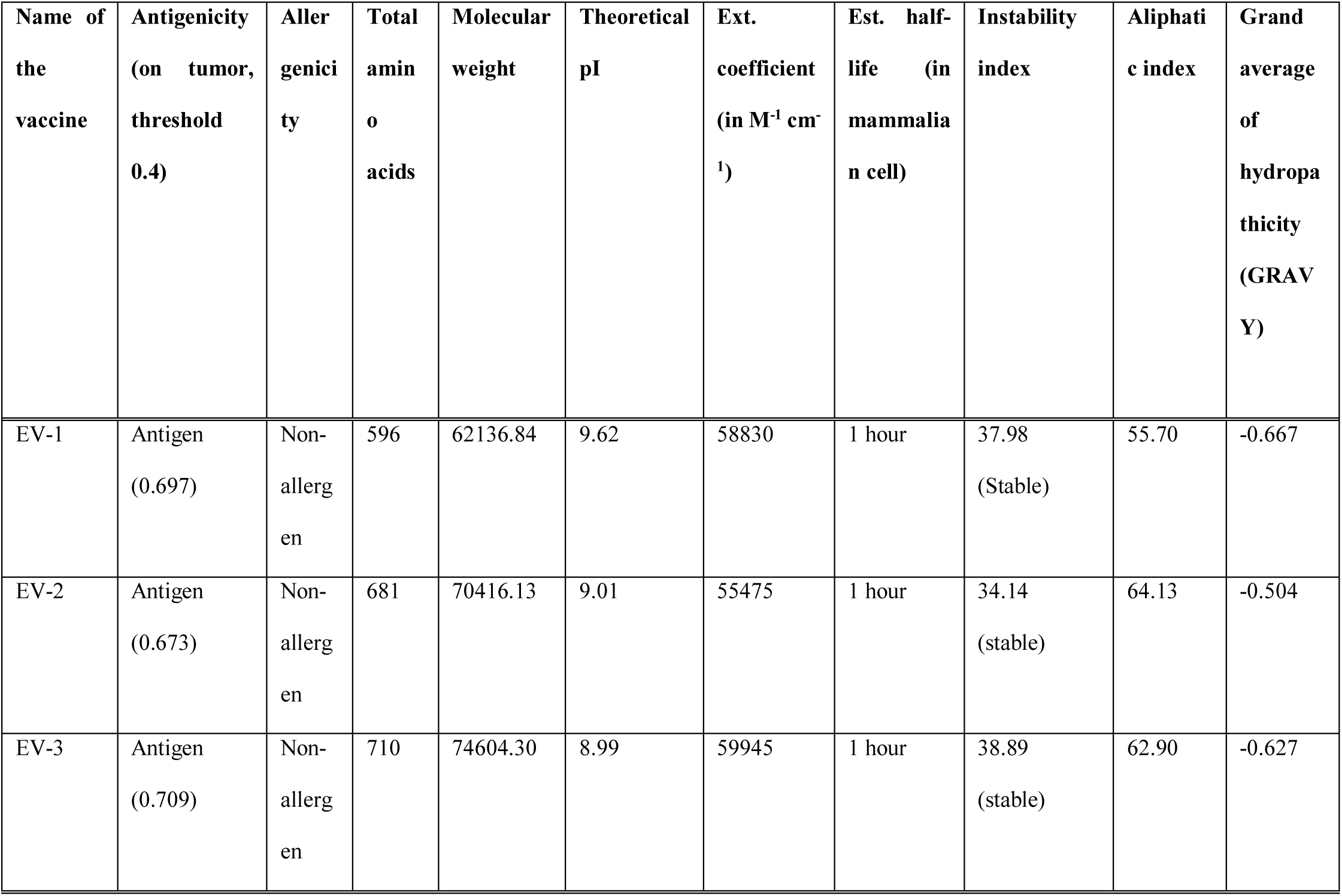
Antigenicity, allergenicity and physicochemical property analysis of the three vaccine constructs.

### 3.9. Secondary and Tertiary Structure Prediction of the Vaccine Constructs

The secondary structures of the three vaccine constructs were generated by two online tools, PRISPRED (http://bioinf.cs.ucl.ac.uk/psipred/) and NetTurnP v1.0 (http://www.cbs.dtu.dk/services/NetTurnP/). From the secondary structure analysis, it was analyzed that, the EV-3 had the highest percentage of the amino acids (35.2%) in the coil formation and EV-2 the highest percentage of amino acids (53.2%) in the beta-strand formation. However, EV-1 had the highest percentage of 14.5% of amino acids in the alpha-helix formation (**Figure 05** and **Table 12**).

**Figure 05.**
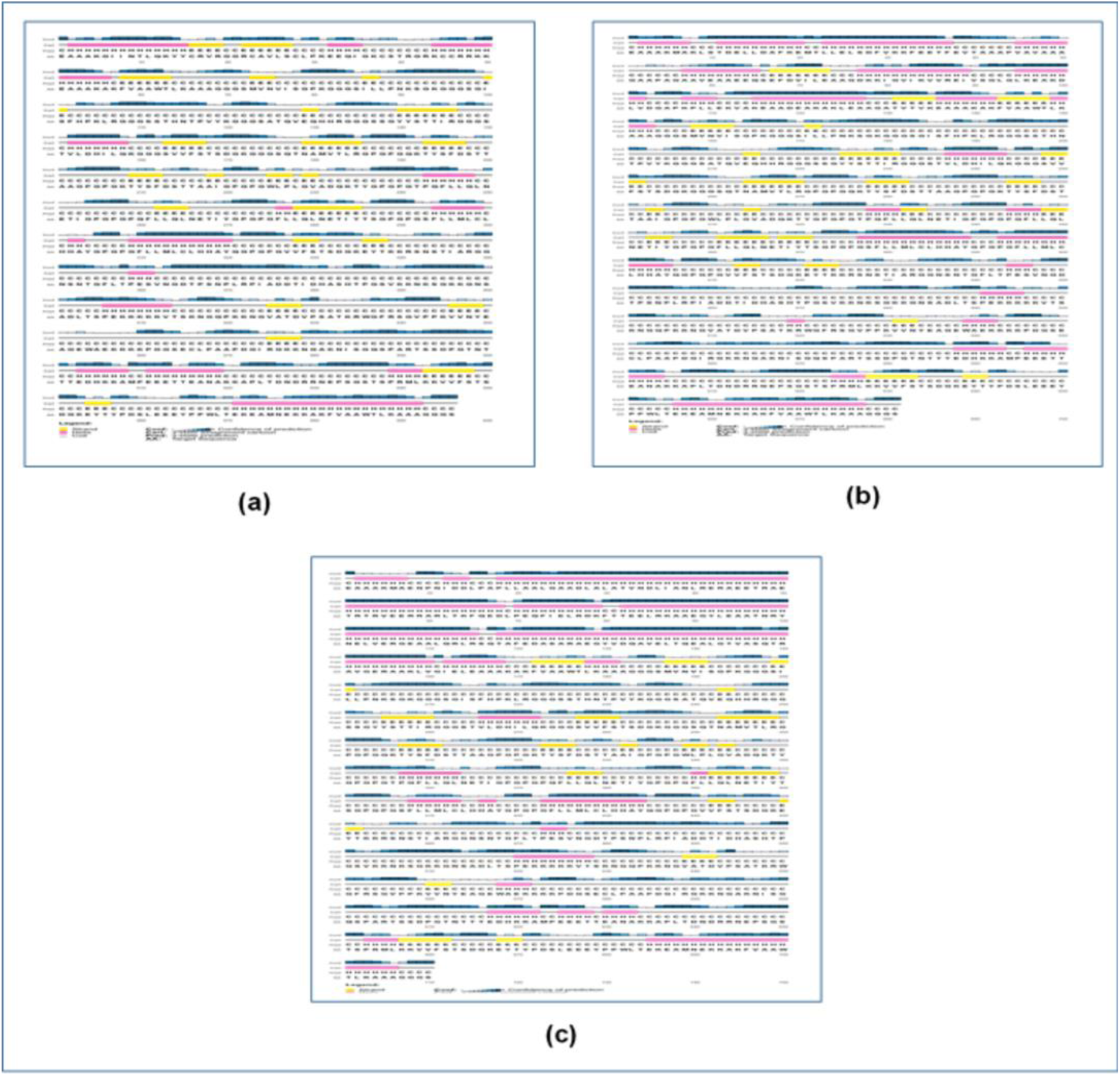
The results of the secondary structure prediction of the constructed HPV-16 vaccines. Here, (a) is EV-1, (b) is EV-2, (c) is EV-3. The secondary structures were predicted using online server PRISPRED (http://bioinf.cs.ucl.ac.uk/psipred/).

**Table 12.**
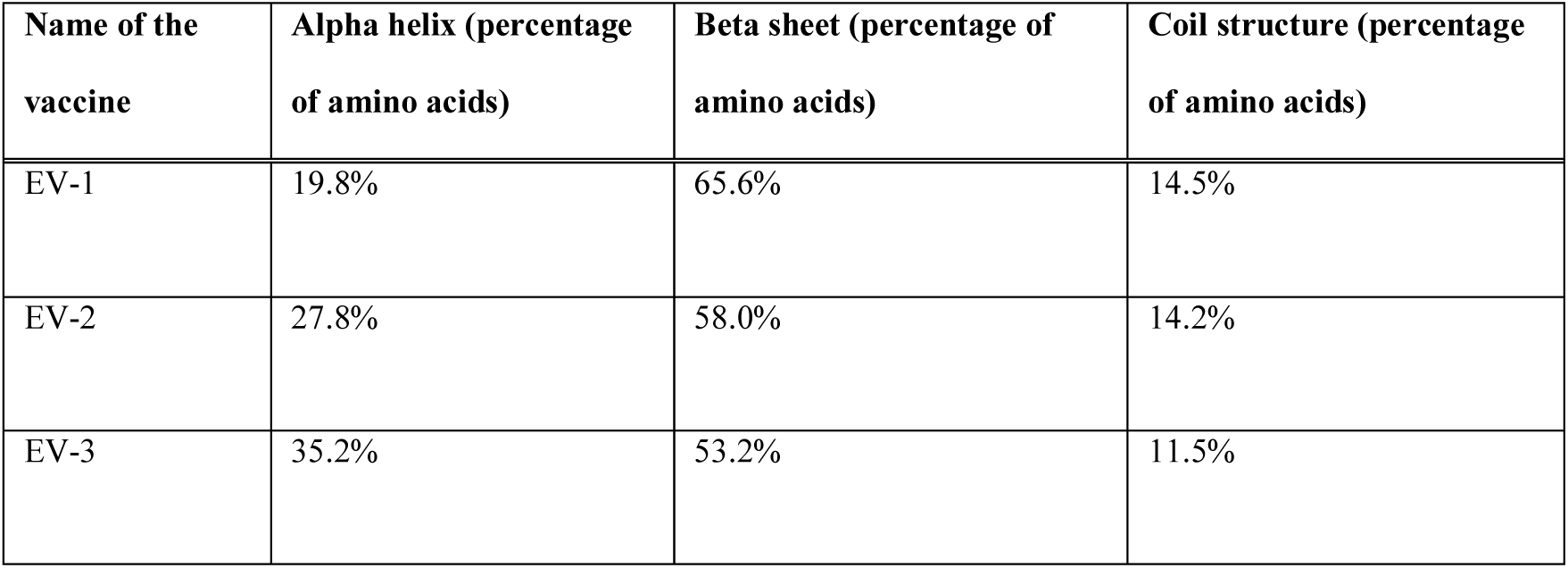
Results of the secondary structure analysis of the vaccine constructs.

The 3D structures of the vaccine constructs were predicted by the online server RaptorX (http://raptorx.uchicago.edu/). All the three vaccines had 5 domains and EV-1 had the lowest p-value of 4.94e-12. The homology modeling of the three dengue vaccine constructs were carried out using 5HJ3C as template from protein data bank (https://www.rcsb.org/). The results of the 3D structure analysis are listed in **Table 13** and illustrated in **Figure 06**.

**Figure 06.**
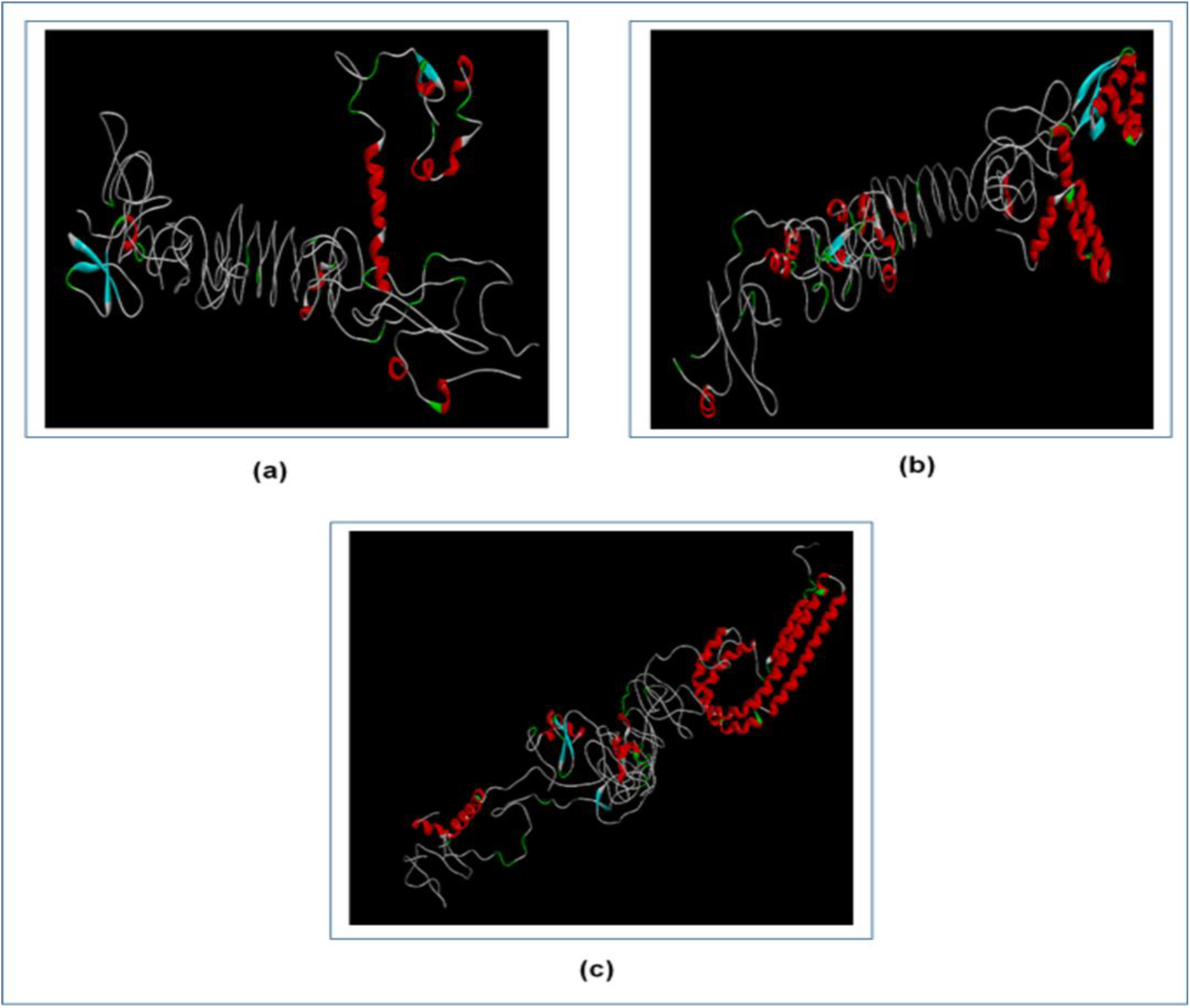
The tertiary structures of the three Ebola vaccines. Here, (a) is the EV-1, (b) is the EV-2 and (c) is the EV-3. The tertiary structures were predicted using the online server tool RaptorX (http://raptorx.uchicago.edu/) and visualized by Discovery Studio Visualizer.

**Table 13.**
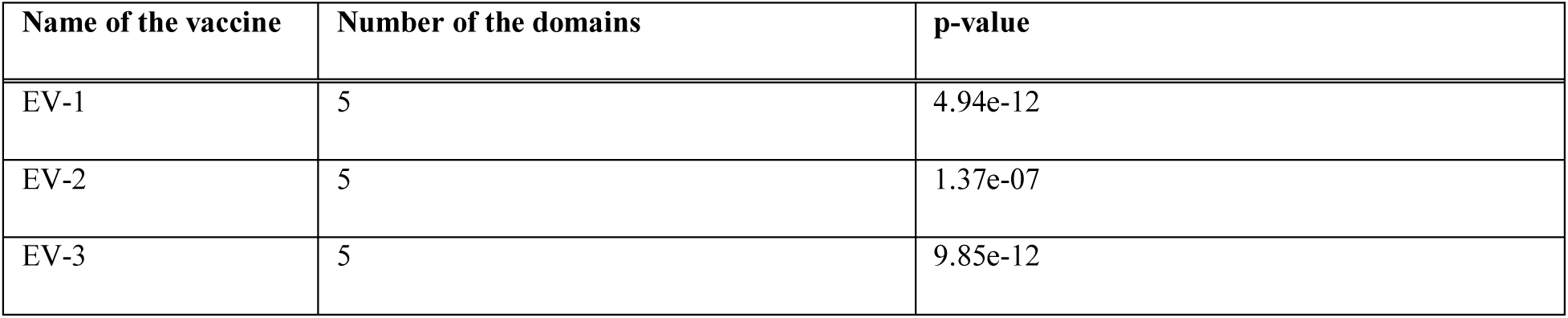
Results of the tertiary structure analysis of the vaccine constructs.

### 3.10. 3D Structure Refinement and Validation

The protein structures generated by the RaptorX server (http://raptorx.uchicago.edu/) were refined using 3Drefine (http://sysbio.rnet.missouri.edu/3Drefine/) and then the refined structures were validated by Ramachandran plot generated by PROCHECK server (https://servicesn.mbi.ucla.edu/PROCHECK/). The analysis showed that EV-1 vaccine had 65.0% of the amino acids in the most favored region, 31.1% of the amino acids in the additional allowed regions, 3.3% of the amino acids in the generously allowed regions and 0.7% of the amino acids in the disallowed regions. The EV-2 vaccine had 66.0% of the amino acids in the most favored region, 29.9% of the amino acids in the additional allowed regions, 3.4% of the amino acids in the generously allowed regions and 0.7% of the amino acids in the disallowed regions. The EV-3 vaccine had 72.1% of the amino acids in the most favored regions, 24.0% of the amino acids in the additional allowed regions, 2.7% of the amino acids in the generously allowed regions and 1.2% of the amino acids in the disallowed regions (**Figure 07**).

**Figure 07.**
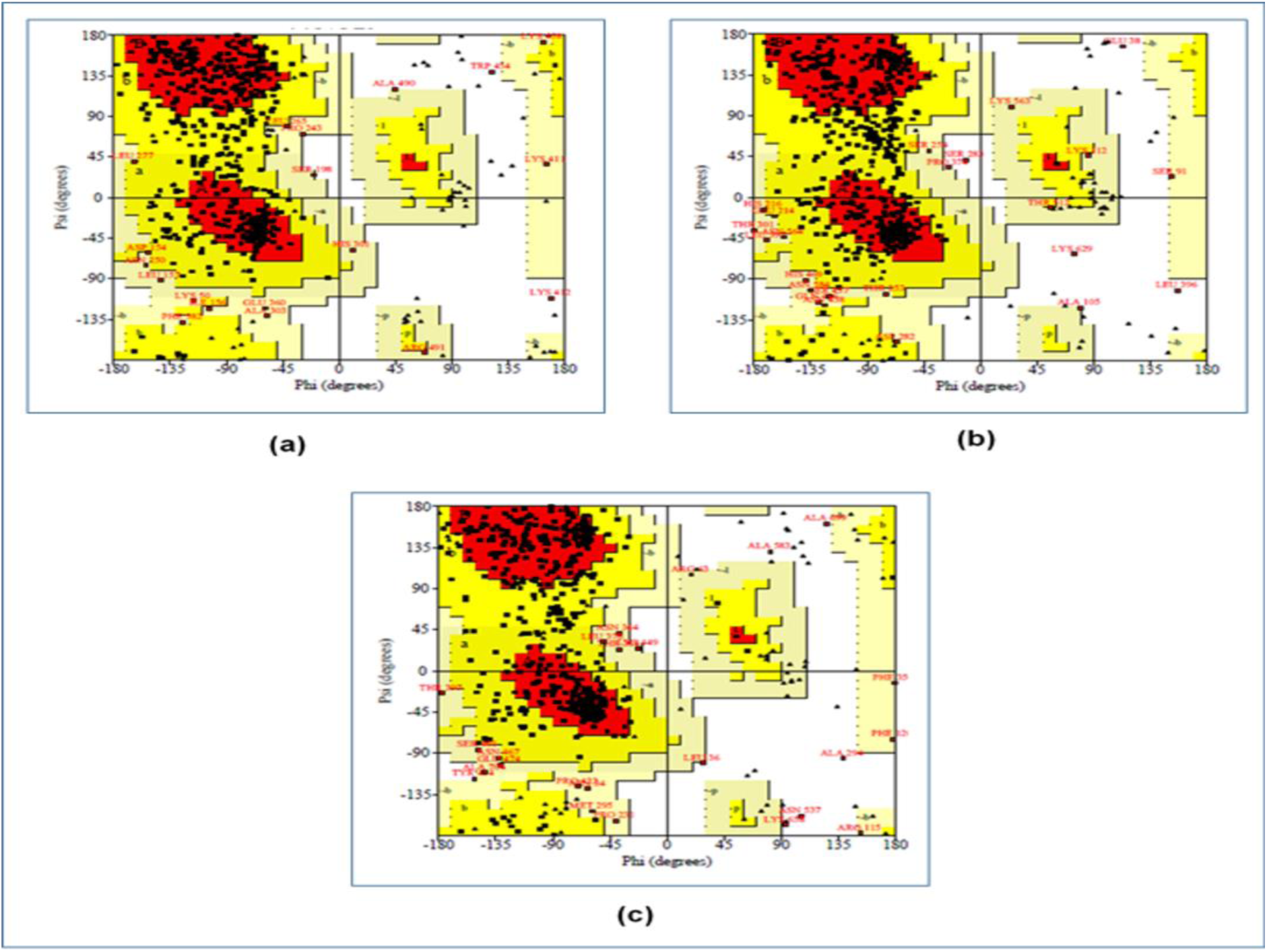
The Ramachandran plot analysis of the three vaccine constructs. (a) EV-1, (b) EV-2, (c) EV-3. The 3D structures of the constructed vaccines were refined using online refinement tool 3Drefine (http://sysbio.rnet.missouri.edu/3Drefine/) and validated by analyzing the Ramachandran plot, generated using the online tool PROCHECK (https://servicesn.mbi.ucla.edu/PROCHECK/).

### 3.11. Protein Disulfide Engineering

In protein disulfide engineering, disulfide bonds were generated for the 3D structures of the vaccine constructs. The DbD2 server identifies the pairs of amino acids that have the capability to form disulfide bonds based on the given selection criteria. In this experiment, we selected only those amino acid pairs that had bond energy value was less than 2.00 kcal/mol. The EV-1 generated 28 amino acid pairs that had the capability to form disulfide bonds. However, only 6 pairs were selected since they had bond energy less than 2.00 kcal/mol: 20 Gly and 127 Gln, 59 Val and 79 Gly, 101 Ser and 114 His, 195 Ser and 238 Pro, 292 Gly and 293 Ser, 318 His and 364 Gly. EV-2 generated 29 pairs of amino acids that had the capability to form disulfide bonds, however, only 4 pairs were selected: 60 Glu and 67 Glu, 68 Phe and Ala, 240 His and 255 Asp, 498 Val and 560 Arg. EV-3 generfated 28 pairs of amino acids capable of forming disulfide bonds and only 5 pairs of the amino acids were selected: 214 Ile and 261 Gly, 229 Asn and 293 Asn, 332 Ala and 344 Val, 438 Gly and 439 Pro, 459 Thr and 476 Val. The selected amino acid pairs formed the mutant version of the original vaccines in the DbD2 server (**Figure 08**).

**Figure 08.**
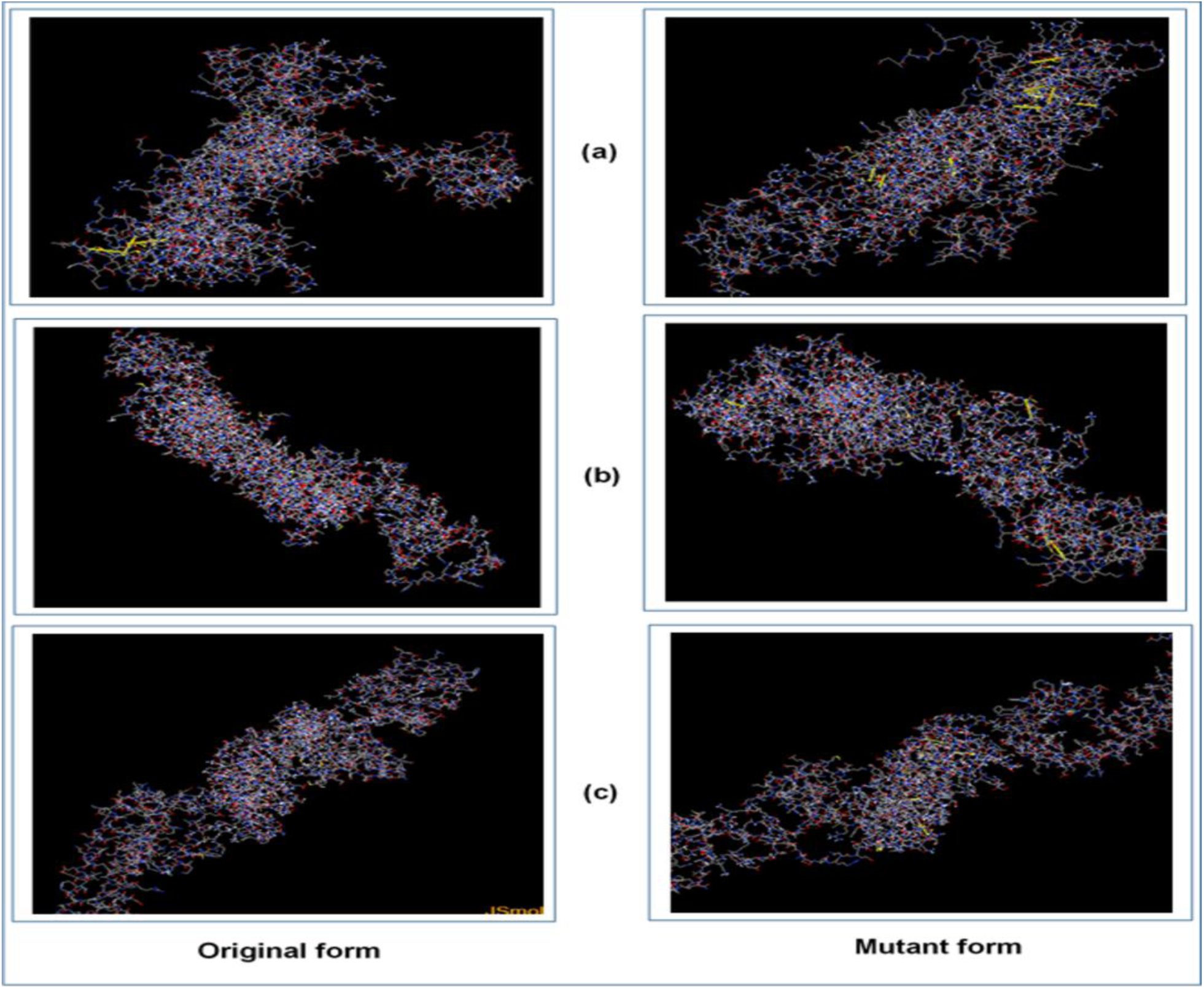
The disulfide engineering of the three vaccine constructs, both the original (left) and mutant (right) forms are shown. Here, (a) EV-1, (b) EV-2, (c) EV-3. The disulfide engineering was conducted using the online tool DbD2 server (http://cptweb.cpt.wayne.edu/DbD2/).

### 3.12. Protein-Protein Docking Study

The protein-protein docking of the vaccine constructs and the MHC alleles were carried out by several online tools for enhancing the accuracy of the prediction: ClusPro 2.0 (https://cluspro.bu.edu/login.php), PatchDock (https://bioinfo3d.cs.tau.ac.il/PatchDock/php.php) and HawkDock server (http://cadd.zju.edu.cn/hawkdock/). The docked complexes that were generated by ClusPro 2.0 and PatchDock were further analyzed by PRODIGY tool of HADDOCK webserver (https://haddock.science.uu.nl/) and FireDock server (http://bioinfo3d.cs.tau.ac.il/FireDock/php.php), respectively. The PRODIGY server generated binding affinity score (in kcal/mol) and FireDock server generated the global energy of the docked complexes. However, HawkDock generated ranking scores along with the binding free energy (in Kcal/mol). The binding free energy was generated after the MM-GBSA study in the HawkDock server. The vaccine constructs were also docked with the TLR-8.

EV-1 showed the best performances in the results generate by the ClusPro 2.0 and PRODIGY server. It had the lowest binding affinity when docked against DRB3*0202 (-17.3 kcal/mol), DRB1*0101 (-19.2 kcal/mol), DRB3*0101 (-19.2 kcal/mol), DRB1*0401 (-21.5 kcal/mol), DRB1*0301 (-17.9 kcal/mol) and TLR-8 (-21.9 kcal/mol). When the docking was performed by the PatchDock server, EV-3 showed the best performances with the lowest score while docking with DRB3*0202 (-19.59), DRB5*0101 (-23.35), DRB3*0101 (-9.89) and TLR-8 (-22.21). However, EV-1 showed the best performances and lowest scores, when docked against all the selected MHC alleles by the HawkDock server and also when analyzed by the MM-GBSA study. For this reason, EV-1 should be considered as the best vaccine construct among the three vaccine constructs, since it generated the best values among the three vaccines in most of the studies (**Figure 09**). **Table 14** lists the results of the docking study of the three Ebola vaccine constructs.

**Figure 09.**
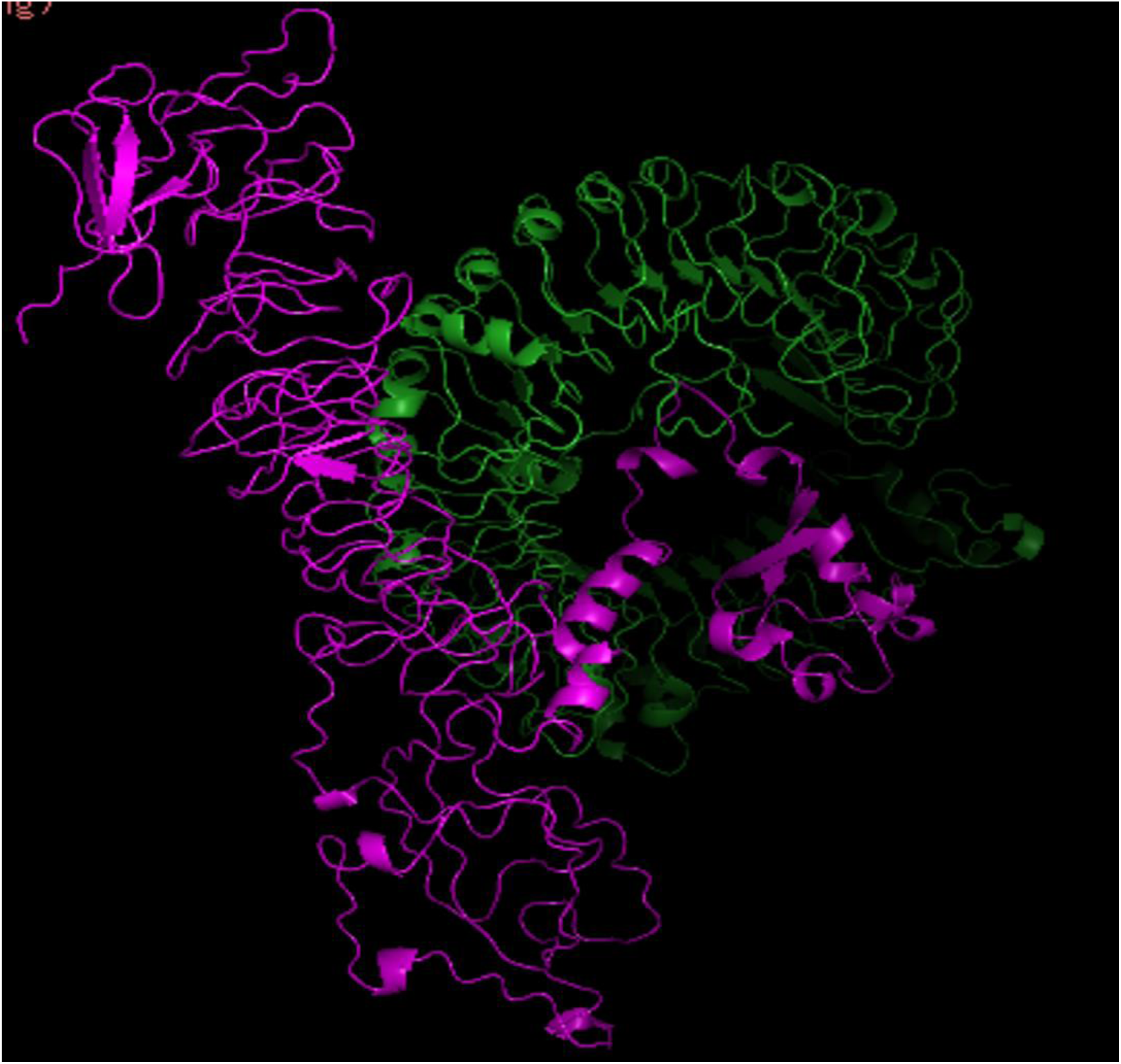
Figure showing the interaction between the ligand protein, EV-1 and receptor protein, TLR-8. The ligand protein is indicated by pink color and the receptor protein is indicated by green color. The visualization of the interactions of the ligands and their receptors were performed by PyMol tool.

**Table 14.**
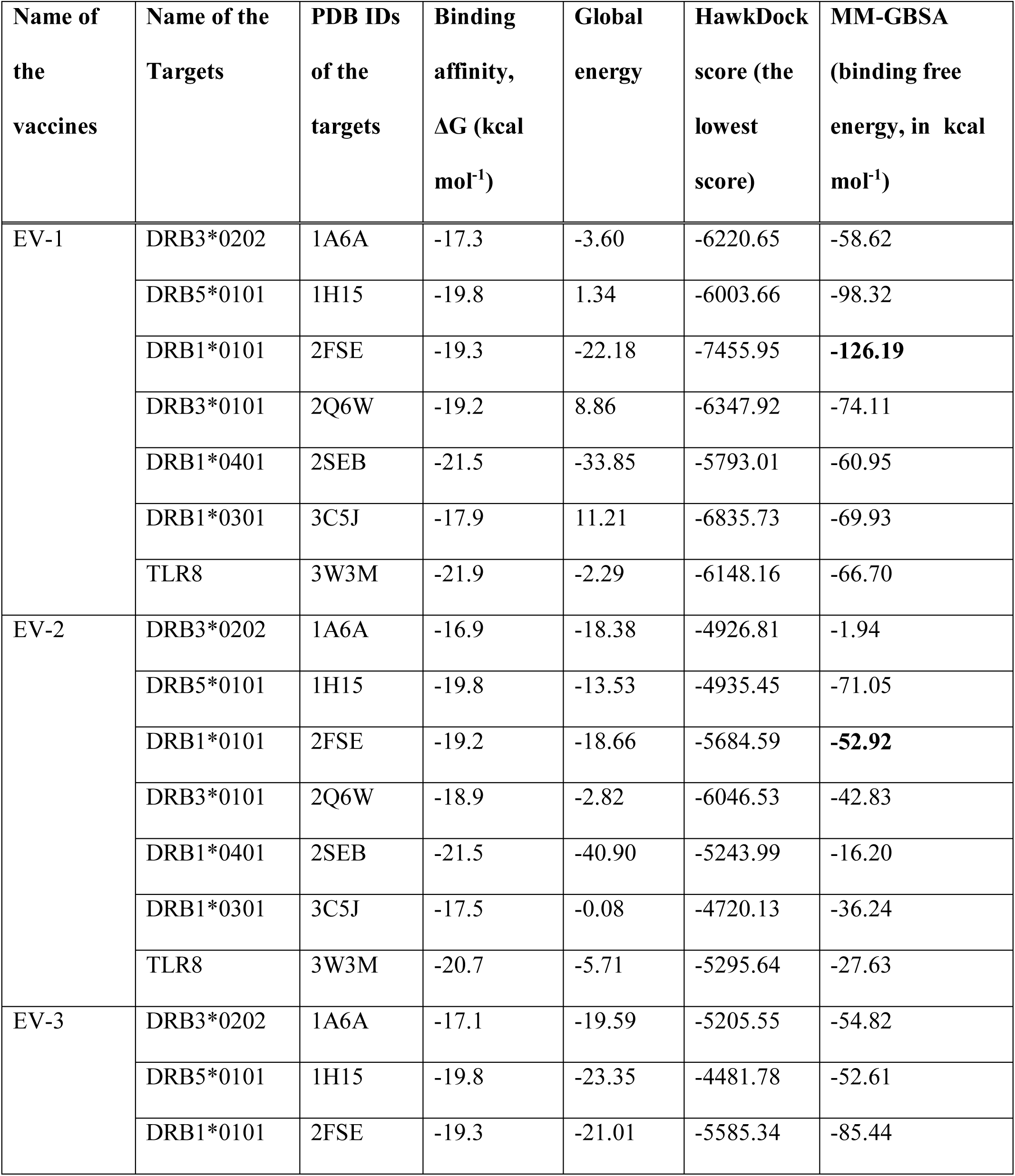
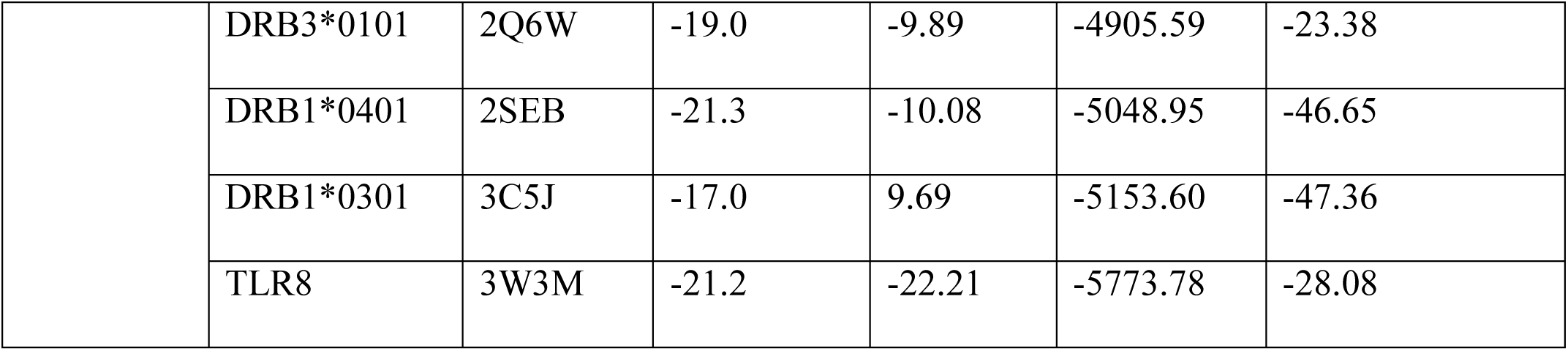
Results of the docking study of all the vaccine constructs.

### 3.13. Molecular Dynamics Simulation

**Figure 10** illustrates the molecular dynamics simulation and normal mode analysis (NMA) of EV-1-TLR-8 docked complex. The deformability graphs of the complex illustrates the peaks in the graphs which represent the regions of the protein with deformability (**Figure 10b**). The B-factor graphs of the complexes give easy understanding and visualization of the comparison between the NMA and the PDB field of the docked complex (**Figure 10c)**. The eigenvalues of the complex is illustrated in **Figure 10d**. EV-1 and TLR8 docked complex generated eigenvalue of 2.129193e-05. The variance graph indicates the individual variance by red colored bars and cumulative variance by green colored bars (**Figure 10e**). **Figure 10f** illustrates the co-variance map of the complex where the correlated motion between a pair of residues are indicated by red color, uncorrelated motion is indicated by white color and anti-correlated motion is indicated by blue color. The elastic map of the complex represents the connection between the atoms and darker gray regions indicate stiffer regions (**Figure 10g**) [67, 68, 69].

### 3.14. Codon Adaptation and In Silico Cloning

For in silico cloning and plasmid construction, the protein sequences of the best selected vaccines were adapted by the JCat server (http://www.jcat.de/).

**Figure 10.**
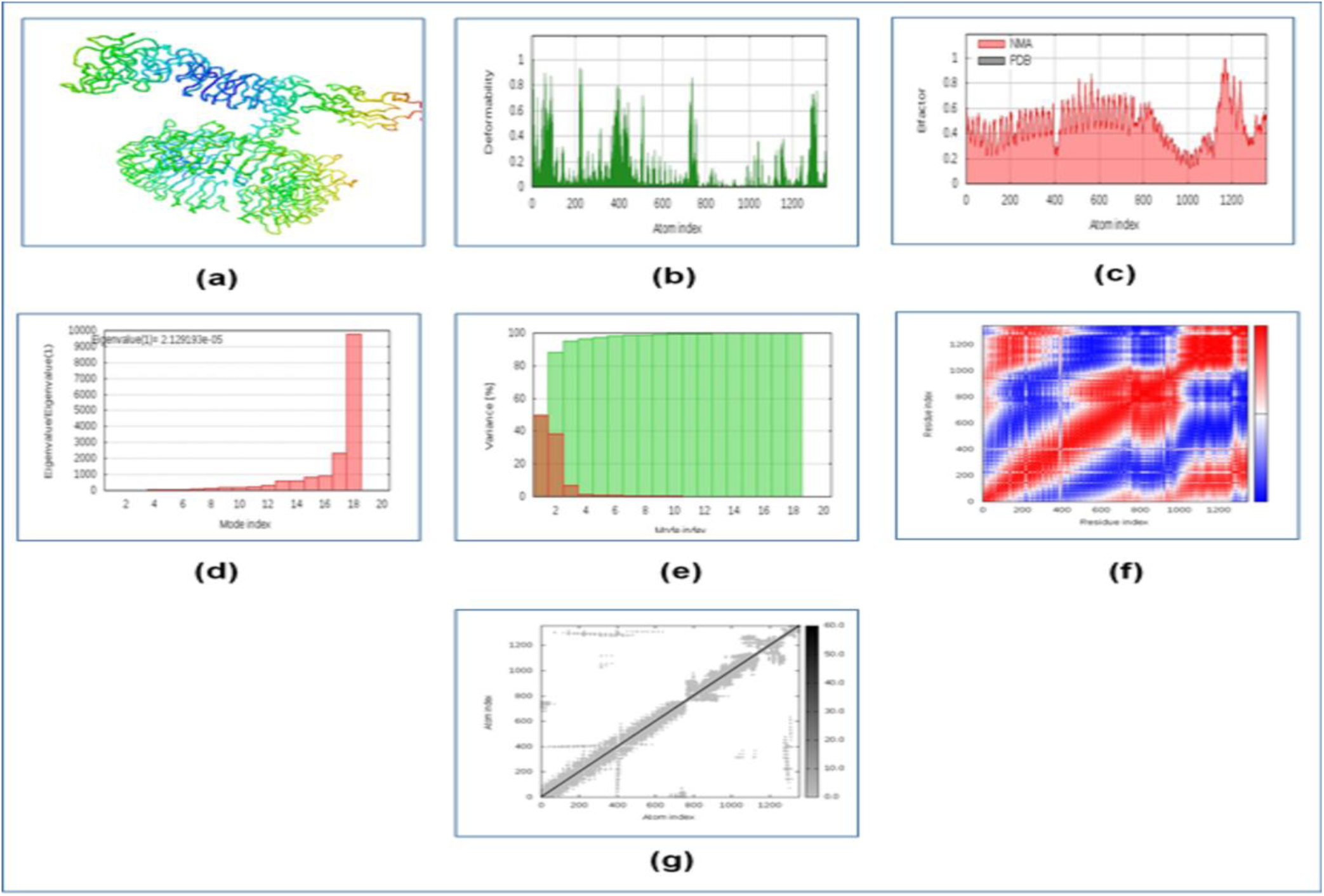
Figure displaying the results of molecular dynamics simulation study of EV-1 and TLR-8 docked complex. Here, (a) NMA mobility, (b) deformability, (c) B-factor, (d) eigenvalues, (e) variance (red color indicates individual variances and green color indicates cumulative variances), (f) co-variance map (correlated (red), uncorrelated (white) or anti-correlated (blue) motions) and (g) elastic network (darker gray regions indicate more stiffer regions).

Since the EV-1 protein had 596 amino acids, after reverse translation, the number nucleotides of the probable DNA sequence of EV-1 would be 1788. The codon adaptation index (CAI) value of 0.943 of EV-1 indicated that the DNA sequences contained higher proportion of the codons that are most likely to be present and used in the cellular machinery of the target organism *E. coli* strain K12 (codon biasness). For this reason, the production of the EV-1 vaccine would be carried out efficiently [73, 74]. The GC content of the improved sequence was 50.73%. The predicted DNA sequence of EV-1 was inserted into the pET-19b vector plasmid between the SgrAI and SphI restriction sites. Since the DNA sequence did not have restriction sites for SgrAI and SphI restriction enzymes, SgrA1 and SphI restriction sites were conjugated at the N-terminal and C-terminal sites, respectively, before inserting the sequence into the plasmid pET-19b vector. The newly constructed cloned plasmid would be 7360 base pair long, including the constructed DNA sequence of the EV-1 vaccine (the EV-1 vaccine DNA sequence also included the SgrAI and SphI restriction sites) (**Figure 11** and **Figure 12**).

**Figure 11.**
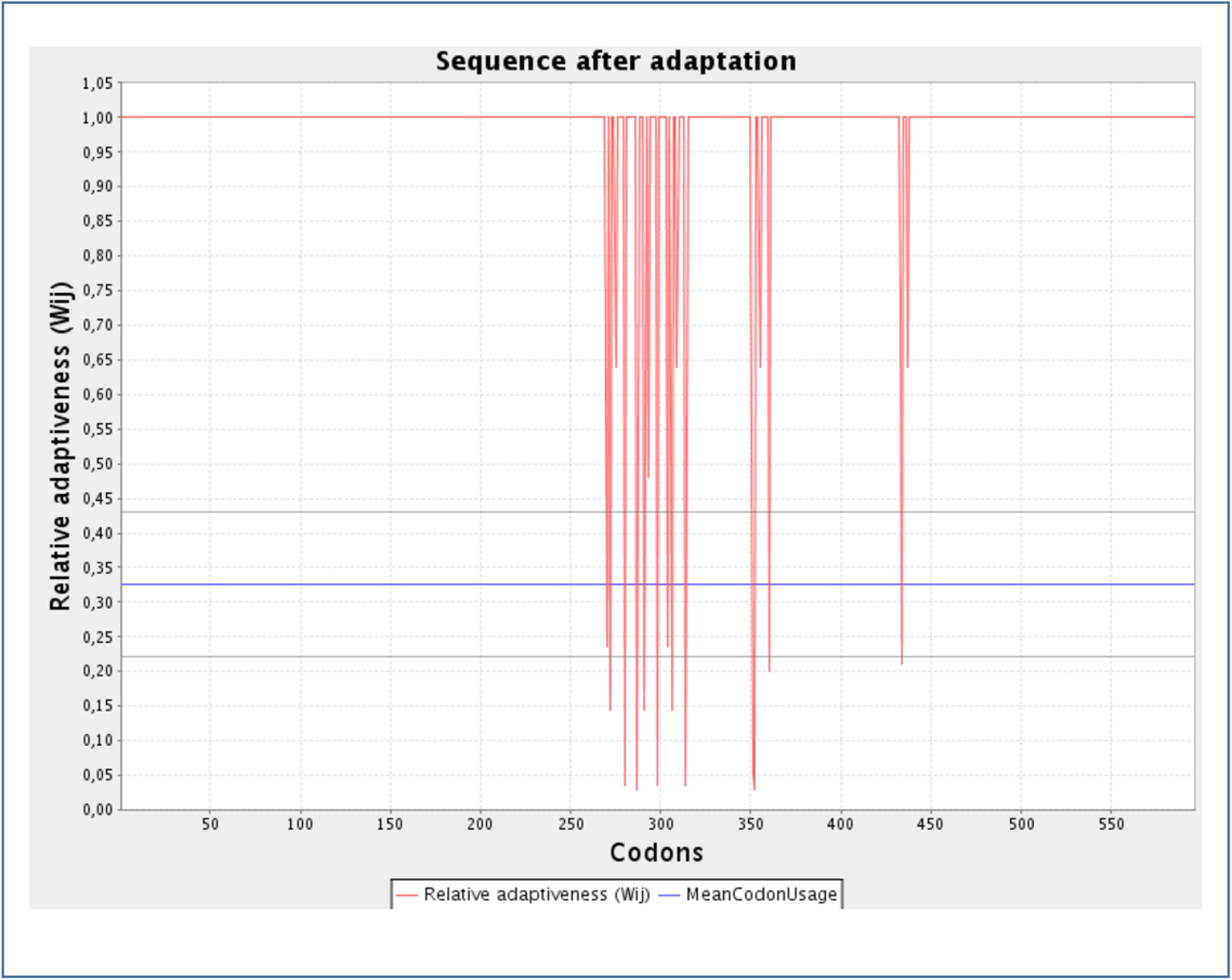
Figure showing the codon adaptation graphs of the EV-1 vaccine. The codon adaptation of the vaccine constructs were carried out using the server JCat (https://jcatbeauty.com/).

**Figure 12.**
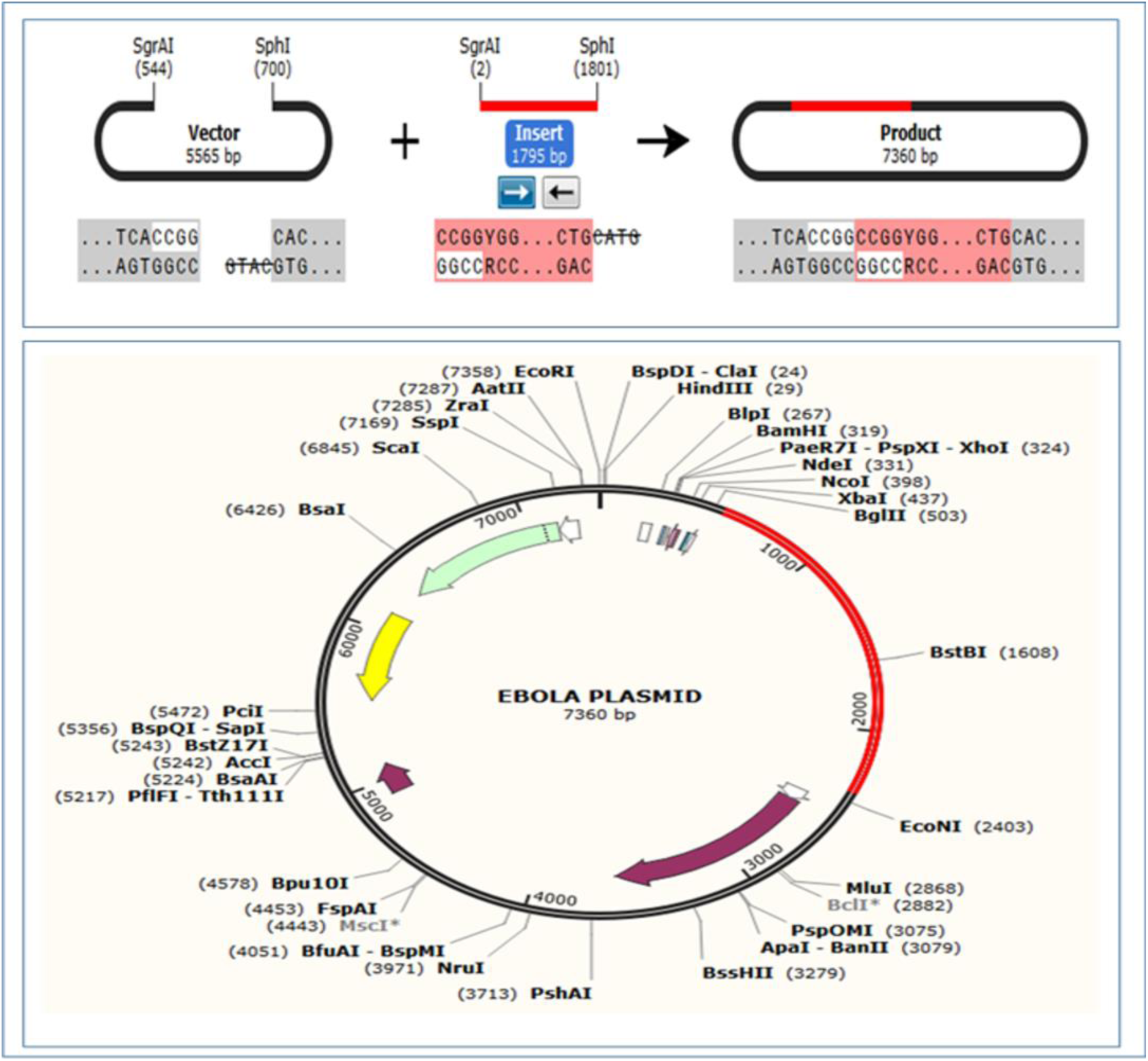
In silico restriction cloning of the EV-1 vaccine sequence in the pET-19b plasmid between the SgrAI and SphI restriction enzyme sites. The red colored marked sites contain the DNA inserts of the vaccines. The cloning was carried out using the SnapGene tool. The two newly constructed plasmids can be inserted into *E. coli* strain K12 for efficient vaccine production.

## 4. Discussion

Vaccine is one of the most important and widely produced pharmaceutical products. However, the development and production of vaccines, is a costly process. Sometimes, it takes many years to develop a proper vaccine candidate against a particular pathogen. In modern times, various methods and tools of bioinformatics, immunoinformatics and reverse vaccinomics are used for vaccine development, which save time and cost of the vaccine development process [75].

The current study was carried out to design possible vaccines against the Zaire Ebola virus, strain-Mayinga 76. The strain of the Ebola virus, against which the vaccines were designed, was selected by reviewing the NCBI database (https://www.ncbi.nlm.nih.gov/). Three proteins of the Zaire Ebola virus strain-Mayinga 76, were selected as the potential targets for the vaccines: matrix protein VP40, envelope glycoprotein and nucleoprotein. The matrix protein VP40 of Ebola virus plays very important roles in the life cycle of a virus, regulating the regulating the viral transcription, virion assembly and budding of viruses from the infected cells [76]. Moreover, the envelope glycoprotein of the Ebola virus mediates the proper viral attachment and entry into the target cell [77]. Studies have also revealed the essential role of nucleoprotein in the replication of Ebola virus [78]. Since these three proteins play very important role in the infection, proliferation and life cycle of the Ebola virus, these proteins could be potential target for vaccine development, thus inhibiting the viral reproduction and infection.

The three protein sequences were retrieved from the UniProt (https://www.uniprot.org/) database. The envelope glycoprotein had accession number: Q05320, matrix protein VP40 had accession number: Q05128 and nucleoprotein had accession number: P18272. Various physicochemical properties like number of amino acids, molecular weight, theoretical pI, extinction co-efficient, instability index, aliphatic index, GRAVY were determined by ProtParam (https://web.expasy.org/protparam/) server. The envelope glycoprotein showed the best performances in the physicochemical property analysis. Envelope glycoprotein is the only stable protein among the three selected proteins of the virus, among the three viral proteins.

However, all the proteins were found to be quite antigenic in the bacteria, parasitic and tumor model, where 0.4 threshold was kept. Matrix protein VP40 generated 0.5692 (bacteria), 0.4598 (tumor), 0.5496 (parasite) in the antigenic test. Envelope glycoprotein showed 0.5852 (bacteria), 0.4933 (tumor), 0.5364 (parasite) scores and nucleoprotein generated 0.5315 (bacteria), 0.5442 (tumor), 0.4692 (parasite) scores in the antigenicity test.

The main cells that function in immunity are the T lymphocytic cell and B lymphocytic cell. After recognized by an antigen presenting cell (like macrophage, dendritic cell etc.), the antigen is presented through the MHC class-II molecule present on the surface of the antigen presenting cell, to the helper T cell. Since, the helper T cell contains CD4+ molecule on its surface, it is also known as CD4+ T cell. After activated by the antigen presenting cell, the T-helper cell then activates the B cell and cause the production of memory B cell and antibody producing plasma B cell. The plasma B cell produce a large number of antibodies and the memory B cell functions as the immunological memory. However, T-helper cell also activates macrophage and CD8+ cytotoxic T cell which destroy the target antigen [79, 80, 81, 82, 83].

The possible T cell and B cell epitopes of the selected viral proteins were determined by the IEDB (https://www.iedb.org/) server. These epitopes should be able to induce strong immune response against the vaccines. The IEDB server generates and ranks the T cell epitopes based on their antigenicity scores (AS) and percentile scores. Based on analyzing AS and percentile scores, ten of the top twenty MHC class-I epitopes or CTL epitopes were selected randomly for further analysis. On the other hand, MHC class-II epitopes or HTL epitopes were also selected based on analyzing the AS or percentile scores. Like the MHC class-I epitopes, ten MHC class-II epitopes were randomly selected from the top twenty epitopes. However, the B cell epitopes that had sequence length of 10 amino acids or over 10 amino acids, were selected for further analysis. The transmemebrane topology of the epitopes were determined to identify whether the epitopes would be present inside or outside of the cell membrane. The transmembrane topology of the selected epitopes were performed using TMHMM v2.0 server (http://www.cbs.dtu.dk/services/TMHMM/).

Antigenicity can be defined as the ability of a foreign substance to act as antigen and activate and stimulate the T cell and B cell responses, through their antigenic determinant portion or epitope [84]. The allergenicity of a substance refers to the ability of that substance to act as allergen and induce potential allergic reactions within the body [85]. When designing a vaccine, epitopes of a protein that remain conserved across various strain, are given much priority than genomic regions that are highly variable because the conserved epitopes of protein(s) provide broader protection across various strains and species [86]. When selecting the best epitopes for vaccine construction, some criteria were maintained: the epitopes should be highly antigenic, so that they can induce high antigenic response, the epitopes should be non-allergenic in nature, for this reason, they would not be able to induce any allergenic reaction in an individual and the epitopes should be non-toxic. The epitopes with 100% conservancy and over 50% minimum identity were selected for vaccine construction, so that, the conserved epitopes would be able to provide protection against various strains. The conservancy analysis was carried out using Zaire Ebola virus strains-Gabon 94 and Kikwit 95, for comparison. Since the selected epitopes were 100% conserved across these strains, for this reason, the vaccine that was constructed for ebola virus strain-Mayinga 76, should provide immunity against the Gabon 94 and Kikwit 95 strains too. Moreover, the cluster analysis of the MHC class-I alleles and MHC class-II alleles were also carried out to determine their relationship with each other and cluster them functionally based on their predicted binding specificity [87].

In the next step, the protein-peptide docking was carried out between the epitopes and the MHC alleles. The MHC class-I epitopes were docked with the MHC class-I allele (PDB ID: 5WJL) and the MHC class-II epitopes were docked with the MHC class-II allele (PDB ID: 5JLZ). The protein-peptide docking was performed to determine the ability of the epitopes to bind with their respective MHC allele. The 3D structures of the selected epitopes were generated by PEP-FOLD3 (http://bioserv.rpbs.univ-paris-diderot.fr/services/PEP-FOLD3/) server. After that, the generated 3D structures were docked with HLA-A*11-01 allele (PDB ID: 5WJL) and HLA DRB1*04-01 (PDB ID: 5JLZ) using PatchDock (https://bioinfo3d.cs.tau.ac.il/PatchDock/php.php) server and refined by FireDock server (http://bioinfo3d.cs.tau.ac.il/FireDock/php.php). GISFHPKLR showed the best result with the lowest global energy of -31.68 among the MHC class-I epitopes of matrix protein VP40. SGYYSTTIR generated the lowest and best global energy score of -42.87 among the MHC class-I epitopes of envelope glycoprotein. QTNAMVTLR generated the best global energy score of -45.40 of the MHC class-I epitopes of nucleoprotein. On the other hand, among the MHC class-II epitopes of matrix protein VP40, KTYSFDSTTAAI generated the best global energy score of -24.32. TPQFLLQLNETI generated the lowest global energy of -2.80 among the MHC class-II epitopes of envelope glycoprotein. Moreover, among the MHC class-II epitopes of nucleoprotein, VVFSTSDGKEYT generated the lowest global energy of -12.06.

After the successful protein-peptide docking, the vaccine construction step was carried out. Three different vaccines were constructed and the three vaccines differed from each other only in their adjuvant sequence. The three vaccines were designated as EV-1, EV-2 and EV-3. EV-1 had beta defensing adjuvant, EV-2 had L7/L12 ribosomal protein as adjuvant and EV-3 had HABA protein as adjuvant. The PADRE sequence was added after the adjuvant sequence and then the CTL, HTL and BCL epitopes were conjugated sequentially. EAAAK, GGGS, GPGPG, KK linkers were used to conjugate the epitopes as well as the adjuvants and PADRE sequence.

After the vaccine construction was performed, the antigenicity, allergenicity and physicochemical property analysis were carried out. All the vaccine constructs were proved to be potential antigenic, however, they were also non-allergenic, for this reason, they should not cause any allergenic reaction within the body. The instability index of a compound refers to the probability of the compound to be stable. IF a compound had instability index of over 40, then the compound is considered to be unstable [88]. The extinction coefficient refers to the amount of light, that is absorbed by a compound at a certain wavelength [89, 90]. EV-3 had the highest extinction co-efficient of 59945 M^-1^ cm^-1^. The aliphatic index of a protein corresponds to the relative volume occupied by the aliphatic amino acids in the side chains like alanine, valine etc. [91]. EV-2 had the highest aliphatic index among the vaccine constructs. Since all the vaccine constructs had instability index of less than 40, for this reason, all of them were considered as stable compounds. All the three vaccine constructs had the similar half-life of 1 hour in the mammalian cell culture and all of them were stable. All of the constructs had quite similar theoretical pI. Quite similar performances were observed by the three vaccine constructs in the physicochemical property analysis.

The secondary structures of the vaccine constructs were generated by two online servers, PRISPRED (http://bioinf.cs.ucl.ac.uk/psipred/) and NetTurnP v1.0 (http://www.cbs.dtu.dk/services/NetTurnP/) and the 3D structures were generated by the server RaptorX (http://raptorx.uchicago.edu/) server. EV-3 had the highest percentage of amino acids in the alpha-helix conformation (35.2%). EV-1 had the highest percentage of amino acids in the beta-sheet conformation with 65.6% of the amino acids in the beta-sheet formation. EV-3 also had the lowest percentage of amino acids (11.5%) in the coil structure of the protein. All the vaccine constructs had 5 domains and EV-2 had the highest p-value of 1.37e-07, determined by the RaptorX server.

In the next step, the 3D structures were refined by online tool 3Drefine (http://sysbio.rnet.missouri.edu/3Drefine/) and validated by PROCHECK (https://servicesn.mbi.ucla.edu/PROCHECK/) server. EV-3 showed the best performances in the validation experiment with 72.1% of the amino acids in the most favored regions and 24.0% of the amino acids in the additional allowed regions. Moreover, EV-3 had 2.7% of the amino acids in the generously allowed regions and 1.2% of the amino acids in the disallowed regions. After validation of the 3D protein structures, the disulfide engineering of the vaccine constructs were performed using Disulfide by Design 2 v12.2 (http://cptweb.cpt.wayne.edu/DbD2/) server and the amino acid pairs with binding energy value less than 2.0 kcal/mol, were picked up for disulfide bond formation.

To find out the best vaccine construct, the protein-protein docking the vaccine constructs and several MHC alleles: DRB1*0101 (PDB ID: 2FSE), DRB3*0202 (PDB ID: 1A6A), DRB5*0101 (PDB ID: 1H15), DRB3*0101 (PDB ID: 2Q6W), DRB1*0401 (PDB ID: 2SEB), and DRB1*0301 (PDB ID: 3C5J) were conducted. The constructed vaccines were also docked against TLR-8 (PDB ID: 3W3M). The aim of the docking experiment is to find out the best vaccine construct among the constructed vaccines. For enhancing the accuracy of our prediction, three different online servers, ClusPro 2.0 (https://cluspro.bu.edu/login.php), PatchDock (https://bioinfo3d.cs.tau.ac.il/PatchDock/php.php) and HawkDock server (http://cadd.zju.edu.cn/hawkdock/) were used for the docking study. The results of the ClusPro 2.0 and PatchDock were refined and analyzed by PRODIGY tool of HADDOCK webserver (https://haddock.science.uu.nl/) and FireDock server (http://bioinfo3d.cs.tau.ac.il/FireDock/php.php), respectively. EV-1 showed the best and excellent performances in the results generated by HADDOCK and HawkDock servers as well as in the MM-GBSA study. For this reason, EV-1 was considered the best vaccine construct which was selected for further analysis.

The molecular dynamics simulation study was carried out using the docked TLR-8 and EV-1 complex using the online tool iMODS (http://imods.chaconlab.org/). The complex showed quite good results in the molecular dynamics simulation study. The study showed that the complex had quite lesser chance of deformability with quite high eigenvalue of 2.129193e-05. The deformability graph (**Figure 10b**) also confirmed that the location of the hinges in the structures were not quite significant and the complex is quite stable with lower degree of deforming for each individual amino acids residue. However, EV-1 had good number of correlated amino acids (marked by red color) and also showed a large number stiffer regions (marked by darker gray color) (**Figure 10f, Figure 10g, Figure 10f** and **Figure 10g**). Nevertheless, it can be concluded that, the complex showed good results in the molecular dynamics simulation study.

Finally, the designed EV-1 vaccine was adapted for in silico cloning. After reverse transcribed, the codon adaptation was carried for cloning in to *E. coli* strain K12. After codon adaptation, the CAI value of 0.943 was obtained, which supported that, the codon adaptation was carried out perfectly and the DNA sequence contained the codons that are most likely used by the *E. coli* cells. The vaccine DNA sequence was inserted into the SgrAI and SphI restriction sites of the pET-19b plasmid vector for efficient expression and production of the desired vaccine, EV-1 in *E. coli* cells. This insertion produced 7360 base pair long plasmid. The plasmid could be used for producing EV-1 by inserting into the *E. coli* cells. However, more in vitro and in vivo studies should be conducted to finally confirm the safety and efficacy of the predicted best considered vaccines. Moreover, since other vaccines also showed quite good results, they can also be tested to finally confirm the best vaccines among the predicted three vaccines.

## 5. Conclusion

Ebola virus disease or EVD is a type of haemorrhagic fever that is caused by the infection of Ebola virus. With the high mortality rate, Ebola can be considered as one of the fatal viruses in the world. However, no vaccines are yet discovered with satisfactory results, to control the outbreak of Ebola. In this study, a possible subunit vaccine to fight against Ebola was designed using various tools of bioinformatics, immunoinformatics and vaccinomics. Bioinformatics and related technologies are heavily used in drug design and development since these technologies reduce the time and cost of drug R&D processes. In this study, first the potential proteins of the viral structure are identified. Next, the potential epitopes were identified through robust processes and these epitopes were used for vaccine construction. Three possible vaccines were constructed and by conducting the docking experiments, one best vaccine construct was identified. Later, the molecular dynamics simulation study and codon adaptation as well as in silico cloning were performed. However, wet lab researches should be carried out on the findings of this experiment to finally confirm the safety, efficacy and potentiality of the vaccine constructs. Hopefully, this research will raise interests among the scientists of the respective field.

## Data Availability

Authors have made all the data generated during experiment available within the manuscript.

## 6. Acknowledgements

Authors are thankful to Swift Integrity Computational Lab, Dhaka, Bangladesh, a virtual platform of young researchers, for providing the tools.

